# The relative strength and timing of innate immune and CD8 T-cell responses underlie the heterogeneous outcomes of SARS-CoV-2 infection

**DOI:** 10.1101/2021.06.15.21258935

**Authors:** Budhaditya Chatterjee, Harshbir Singh Sandhu, Narendra M. Dixit

**Author notes:** These authors contributed equally (Budhaditya Chatterjee, Harshbir Singh Sandhu).

## Abstract

SARS-CoV-2 infection results in highly heterogeneous outcomes, from cure without symptoms to acute respiratory distress and death. While immunological correlates of disease severity have been identified, how they act together to determine the outcomes is unknown. Here, using a new mathematical model of within-host SARS-CoV-2 infection, we analyze diverse clinical datasets and predict that a subtle interplay between innate and CD8 T-cell responses underlies disease heterogeneity. Our model considers essential features of these immune arms and immunopathology from cytokines and effector cells. Model predictions provided excellent fits to patient data and, by varying the strength and timing of the immune arms, quantitatively recapitulated viral load changes in mild, moderate, and severe disease, and death. Additionally, they explained several confounding observations, including viral recrudescence after symptom loss, prolonged viral positivity before cure, and mortality despite declining viral loads. Together, a robust conceptual understanding of COVID-19 outcomes emerges, bearing implications for interventions.

**Teaser:** Modeling explains how a subtle interplay between innate immune and CD8 T-cell responses determines the severity of COVID-19.

## Introduction

Coronavirus disease 2019 (COVID-19), a respiratory infection caused by the severe acute respiratory syndrome coronavirus 2 (SARS-CoV-2), evokes remarkably heterogeneous clinical outcomes (1, 2). While some individuals are cured without any symptoms, others suffer mild to moderate symptoms, and yet others experience severe disease requiring hospitalization and intensive care, with a sizeable fraction of the latter suffering death (1–3). Several demographic correlates of disease severity, such as gender, co-morbidities, and age, have been identified (4). Further, immunological correlates of severe disease outcomes, such as a subdued early innate immune response (5), and a late surge of proinflammatory cytokines (6, 7) have also been reported. Yet, what determines this diversity of outcomes has remained an outstanding question, challenging our understanding of infectious disease biology and, more immediately, precluding optimal strategies for combating the raging COVID-19 pandemic.

While viral factors, including emerging mutations (8), may have a role in determining the outcomes, the heterogeneous outcomes were reported in early studies (2, 3), before the majority of the clades of SARS-CoV-2 emerged (9), suggesting that the heterogeneity potentially originates from the variability in the host immune responses to the infection (6). Rapidly accumulating evidence reinforces the role of the immune response, particularly of the innate and the CD8 T-cell responses, in determining disease outcomes: Soon after infection, an innate immune response is first mounted, involving the production of cytokines, particularly type I and type III interferons, by virus-infected and immune cells (10). Interferons work across viruses and, through autocrine and paracrine signaling mechanisms, can reduce viral production from infected cells and render proximal target cells temporarily resistant to infection, controlling disease progression (10, 11). With SARS-CoV-2, patients with mild disease had higher levels of interferons in their upper respiratory airways than those with more severe disease, suggesting that robust innate immune responses contribute to reduced severity of infection (5).

A few days into the infection, the adaptive immune response involving virus-specific effector CD8 T-cells is triggered. CD8 T-cells are thought to play a critical role in the clearance of SARS-CoV-2 (7): The earlier the first detectable CD8 T-cell response, the shorter is the duration of the infection (12). CD8 T-cell numbers were higher in the bronchoalveolar lavage fluids of individuals with mild/moderate symptoms than in those with severe infection (13). Clonal expansion of CD8 T-cells was compromised in patients with severe symptoms (13, 14). Infected individuals often suffer lymphopenia (15, 16), with the extent of lymphopenia correlated with disease severity (15, 17). Finally, the severity of the symptoms was proportional to the level of exhaustion of CD8 T-cells (15, 17). Accordingly, a combination of the innate and CD8 T-cell responses appears to drive viral clearance.

Once the disease is resolved, typically in 2-3 weeks, the cytokines and activated CD8 T-cell populations decline and eventually fade away, leaving behind memory CD8 T-cells (7). If the disease is not resolved in a timely manner, uncontrolled cytokine secretion may result, triggering immunopathology and severe disease (6). Indeed, an elevated interferon response was detected in the lower respiratory tracts of severely infected and deceased patients (5, 18, 19), with the lung suffering the most damage (20). Innate immune cell-types, such as neutrophils, macrophages and natural killer cells, which are thought not to contribute significantly to clearance, may nonetheless worsen the damage (6, 21). Prolonged disease, where viral load could be detected in patients over extended durations–up to 66 days on average in some cohorts–has been reported (22–24). Proliferation and differentiation of CD8 T-cells were compromised in prolonged SARS-CoV-2 positive patients (22). The innate immune and CD8 T-cell responses thus appear to be involved in these undesirable outcomes of the infection as well.

Antibodies, the other component of the adaptive immune response, arise much later, a couple of weeks into the infection (7, 25). While important in vaccine-mediated protection (26, 27), their role in clearing infection in the unvaccinated is thought to be less significant than that of CD8 T-cells (7). Antibody titers are higher in severely infected than in mildly infected individuals (7). Whereas a subset of antiviral antibodies possibly contribute to the clearance of infection (28), autoantibodies, typically generated in COVID-19 patients, against cytokines and cell surface and structural proteins of the host, may adversely affect clinical outcomes (29).

Based on these observations, we hypothesized that the strength and the timing of the innate and the CD8 T-cell responses were the predominant factors responsible for the heterogeneous outcomes of SARS-CoV-2 infection. To test this hypothesis, we developed a mathematical model of within-host SARS-CoV-2 dynamics that incorporated the key features of the innate and the CD8 T-cell responses. We validated the model against patient data and employed it to elucidate the interplay of the two immune arms in the outcomes realized.

## Results

### Mathematical model of within-host SARS-CoV-2 dynamics

We considered an individual infected by SARS-CoV-2. We modeled disease progression in the individual by following the time-evolution of the population of infected cells (*I*), the strength of the effector CD8 T-cells (*E*), the cytokine-mediated innate immune response (*X*), and tissue damage (*D*) (Figure 1). We considered the essential interactions between these entities (30) and constructed the following equations to describe their time-evolution:

**Figure 1:**
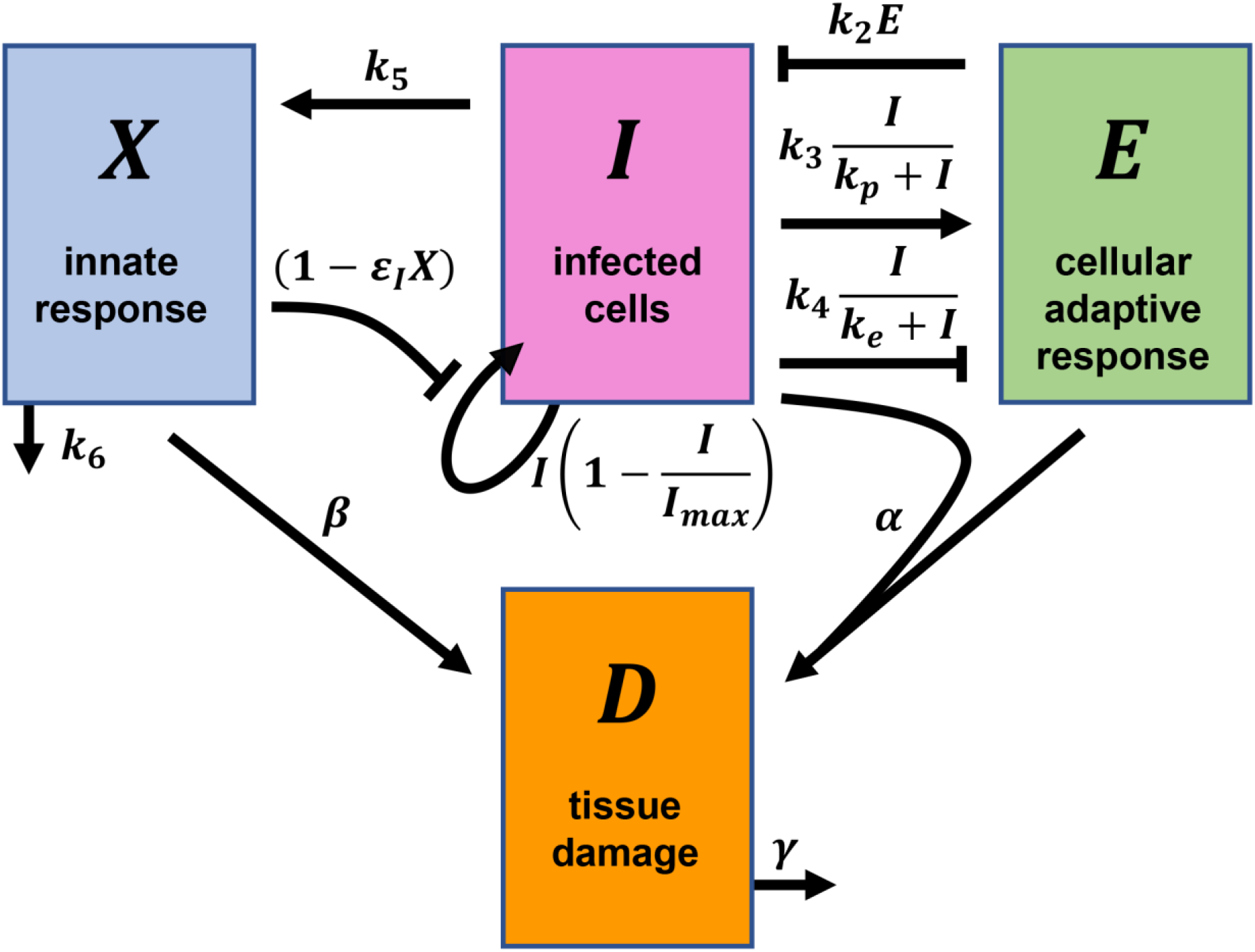
Schematic of the mathematical model of within-host SARS-CoV-2 infection. *I* represents infected cells, *X* represents the innate immune response, *E* the CD8 T-cell response and *D* the tissue damage. Arrows and blunt-head arrows depict positive and negative regulation, respectively. The parameters and expressions shown next to the arrows are described in the text.

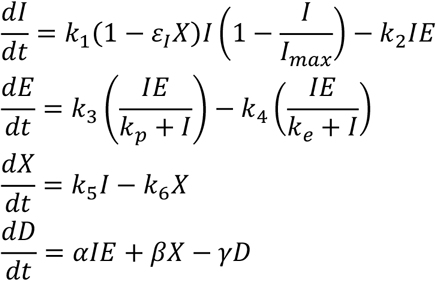

Here, the infected cells follow logistic growth (30), with a per capita growth rate *k*_1_ and carrying capacity *I*_*max*_. This growth represents the infection of target cells by virions produced by infected cells (30). *I*_*max*_ is the maximum number of cells that can get infected, due to target cell or other limitations. The growth rate *k*_1_ is assumed to be reduced by the innate immune response, *X*, with the efficacy *ε*_*I*_*X*, due to interferon-mediated protection of target cells and/or lowering of viral production from infected cells (10). Effector cell-mediated killing of infected cells is captured by a mass action term with the second-order rate constant *k*_2_. The proliferation and exhaustion of CD8 T-cells are both triggered by infected cells at maximal per capita rates *k*_3_ and *k*_4_, respectively. *k*_*p*_ and *k*_*e*_ are the levels of infected cells at which the proliferation and exhaustion rates are half-maximal, respectively. Following previous studies, we let *k*_3_ < *k*_4_ and *k*_*p*_ < *k*_*e*_, so that proliferation dominates at low antigen levels and exhaustion at high antigen levels (30–32). The innate response, *X*, is triggered by infected cells at the per capita rate *k*_5_ and is depleted with the first-order rate constant *k*_6_. To assess the severity of infection, we employ *D*, which represents the instantaneous tissue damage, with contributions from CD8 T-cell mediated killing of infected cells, determined by *αIE*, and from proinflammatory cytokines, represented by *βX*. Inflamed tissue may recover with the first order rate constant *γ*.

Solving these equations would predict the time-course of the infection. We tested the model by applying it to describe available patient data of viral load changes.

### Model fits patient data

A number of studies have reported viral load measurements during the course of SARS-CoV-2 infection (33, 34). In most studies, measurements begin from the time of symptom onset because the time of contracting the disease is rarely known. Because the prodromal period may vary substantially across individuals (35), measurements from symptom onset may miss the initial phases of the immune response, which can be an important determinant of disease outcome. In asymptomatic individuals, this early response clears the infection (36). We therefore sought datasets that included accurate estimates of the time of contracting the disease. Fortunately, we found such data in a study of one of the first SARS-CoV-2 transmission chains in Germany in early 2020 (37, 38). The study traced the dates of first exposure to the virus for each patient in the transmission chain (38) (supplementary text section A; supplementary table S1). Further, daily viral load data, measured in nasopharyngeal swab and sputum samples, for all patients starting from the onset of symptoms or earlier were reported (37). Data from the nasopharyngeal swabs are thought not to be the best correlates of disease outcome and severity (39). The sensitivity of SARS-CoV-2 detection in sputum is substantially higher than in nasopharyngeal swabs (40). We therefore employed data from the sputum samples in this study. We considered data from day zero to day 15 into the infection (supplementary text section A; supplementary tables S1-S3). Beyond two weeks, the humoral response is mounted in most patients (7, 25), the role of which, as mentioned above, is poorly understood (7).

We fit our model to the above viral load data, representing the dynamics of the infection and immune responses in the respiratory tract. All patients in this dataset had mild symptoms, which waned by day 7 after the first virological test. The patients were of working age and otherwise healthy. In such patients, markers of T-cell exhaustion are not significantly higher than healthy individuals and are markedly lower than severely infected patients (15). Therefore, to facilitate more robust parameter estimation, we ignored CD8 T-cell exhaustion in the present fits (by fixing *k*_4_ = 0). Furthermore, we assumed that the viral population, *V*, is in a pseudo-steady state with the infected cell population, so that *V* ∝ *I*. Since, the dynamics of tissue damage (*D*) is dependent on but does not affect the dynamics of infected cells (*I*), CD8 T-cells (*E*) and the cytokine mediated innate response (*X*), in our model, we ignored *D* for the present fitting. This is further justified because the patients considered for fitting are mildly/moderately infected, and are expected to suffer minimal tissue damage. Because the patients were all similar, we assumed that *I*_*max*_ would be similar in them and proportional to *V*_*max*_, the highest viral load reported across the patients. We thus fit log_10_(*I*/*I*_*max*_) calculated with our model to the normalized data of log_10_(*V*/*V*_*max*_). Our fits were not sensitive to *I*_*max*_ (supplementary tables S4, S5). We allowed a delay following exposure to account for the incubation period before viral replication can begin. We used a nonlinear mixed-effects modelling approach for parameter estimation (41). Our model provided good fits to the data (figure 2, first column of subplots) and yielded estimates of the parameters at the population-level (supplementary table S6) and for the individual patients (supplementary table S7).

**Figure 2:**
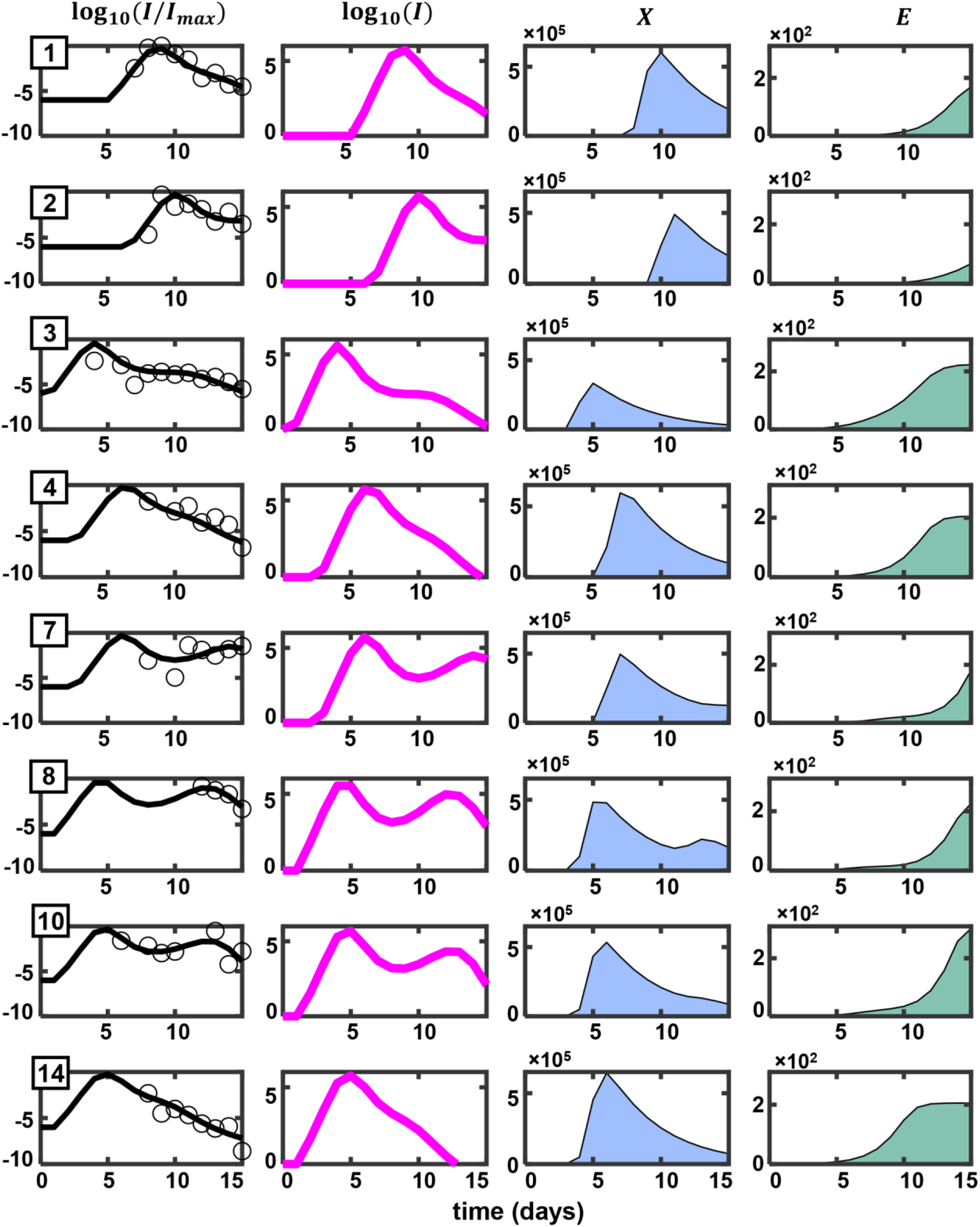
Fits of the mathematical model to patient data. X-axis represents time from viral exposure in all subplots. The quantity plotted on the Y-axis for all subplots in a given column is mentioned at the top of the column. The first column shows data from patients in open circles (37). These represent normalized viral loads from sputum samples. Best-fit model predictions are shown as black curves. Patient IDs as provided in Böhmer et al. (38) are in the top-left corner of each subplot. The magenta curves in the second column, the blue area plots in the third column and the green area plots in the fourth column represent the corresponding dynamics of infected cells, cytokine mediated innate immune response and effector CD8 T-cell mediated adaptive immune response, respectively. Parameter values used are listed in supplementary table S7.

To ascertain the robustness of our model and fits, we tested several variants of our model. We fit variants without the adaptive response, without the innate response, with a logistic growth formulation of the innate immune response, with the innate response amplifying the adaptive response, or combinations thereof to the same data (supplementary text section B, supplementary table S8). The fits were all poorer than the present model (figure 1, supplementary table S8). We thus employed our present model for further analysis.

### Model elucidates distinct patterns of viral clearance and associated immune responses

The best-fits above yielded important insights into the underlying dynamics of disease progression and clearance. First, the time between viral exposure and noticeable escalation of the viral load, i.e., the post-exposure delay in viral replication, varied from 0.8 d to 6.6 d in the patients analyzed, with a mean of 2.7±0.8 days, reflecting the variability in the time of the establishment of systemic infection following exposure, and consistent with the variable prodromal period observed (35). The initial, possibly stochastic (42), events during the establishment of infection might be associated with the variability in the delay in viral replication. Second, our model offered an explanation of the two distinct patterns of clearance observed in the patients. Patients 1, 2, 3, 4, and 14 had a single peak in viral load (or infected cell numbers) followed by a decline of viral load leading to clearance (figure 2, second column of subplots). Patients 7, 8 and 10, in contrast, had a second peak following the first. Our model predicted these distinct patterns as arising from the temporal variation in the dynamics of the innate and CD8 T-cell responses.

The interactions between the innate response, *X*, and infected cells, *I*, in our model have signatures of the classic predator-prey system (43) with *I* the prey and *X* the predator: In the absence of *X, I* grows. *I* also triggers *X*, which in turn suppresses *I. X* declines in the absence of *I*. These interactions, as with the predator-prey system (43), predict oscillatory dynamics. Thus, following infection, *I* grows, causing a rise of *X* in its wake. When *X* rises sufficiently, it suppresses *I*. When *I* declines substantially, the production of *X* is diminished and *X* declines. This allows *I* to rise again and the cycle repeats. This cycle is broken in our model by CD8 T-cells, *E*. Viral clearance is not possible in our model without *E* (supplementary figure S1). When *E* rises, it can suppress *I* independently of *X*, breaking the cycle and allowing *X* to dominate *I*. Together, *X* and *E* can then clear the infection. In patients 7, 8, and 10, our best-fits predicted an early innate immune response and a delayed CD8 T-cell response. The second peak was thus predicted as a result of the above predator-prey oscillations that occurred before the CD8 T-cell response was mounted. In patients 3, 4, and 14, a relatively early CD8 T-cell response was predicted, which precluded the second peak. In patients 1 and 2, both the innate and CD8 T-cell responses were delayed, leaving little time for the oscillations to arise in the 15 d period of our observations. We note that interpretations of the multiple peaks in longitudinal viral load data have not been forthcoming (44). Our predictions offer a plausible interpretation.

Third, the transient but robust innate immune response predicted (figure 2, third column of subplots) is consistent with observations in mildly infected patients (45). Fourth, our prediction of the dynamics of the CD8 T-cell response, where a gradual build-up is followed by a stationary phase (figure 2, fourth column) is also consistent with observations. In mildly infected patients, SARS-CoV-2 specific T-cells were detected as early as 2-5 days post symptom onset (12). This effector population remained stable or increased for several months after clinical recovery (16, 46).

Our model thus fit the dynamics of infection in individuals showing mild symptoms and offered explanations of disease progression patterns that had remained confounding. We examined next whether the model could also describe more severely infected patients. For this, we varied different parameters in our model and assessed the resulting dynamical features of the infection.

### Interplay between innate and CD8 T-cell responses underlies heterogeneous disease outcomes

We reintroduced the CD8 T-cell exhaustion term, which we had ignored in the fits above because the patients were mildly infected, and selected associated parameter estimates from previous studies (30). We ensured that this did not affect our fits above (supplementary figure S2).

Next, to estimate the severity of the disease, we also examined the dynamics of the instantaneous tissue damage (*D*). Typically, *D* rose as the infection progressed and declined as it got resolved (supplementary figure S3A, C). We reasoned that the severity of infection would be determined by the maximum tissue damage suffered and the duration for which such damage lasted. Significant damage that is short-lived or minimal damage that is long-lived may both be tolerable and lead to mild symptoms. We therefore calculated the area under the curve (AUC) of *D*, starting from when *D* ascended above its half-maximal level to the time when it descended below that level (supplementary figure S3A), as a measure of immunopathology (*P*) and associated disease severity. (Note that the parameters *α, β*, and *γ*, which describe the dynamics of tissue damage, are unknown constants; our results were not sensitive to their values because changing them only minimally affected the relative extents of immunopathology across different disease severity categories (supplementary text section C; supplementary figure S5).)

With this framework, we varied the strengths of the CD8 T-cell and innate responses, by changing the values of the parameters *k*_3_ and *k*_5_, respectively, and examined the predicted dynamical features (figure 3A). Recall that *k*_3_ is the proliferation rate of CD8 T-cells and *k*_5_ is the rate of generation of the innate immune response. The other parameters were fixed (supplementary table S6) at their population estimates, for which the model elicited clearance of the infection by day 14 (figure 3A, center, subplot with an arrowhead). Increasing *k*_5_ resulted in a decrease in the peak of infected cells (figure 3A, the row of subplots with arrowhead, right to left). With decrease in *k*_5_, the induction of the cytokine mediated antiviral innate response was substantially delayed and that corresponded to an increased number of infected cells (supplementary figure S4A, B). Clearance was achieved in all cases without substantial variation in the infection duration because of the CD8 T-cell response (figure 3A, the row of subplots with arrowhead, right to left). Decreasing *k*_3_ weakened the CD8 T-cell response and increased the duration of the infection (figure 3A, the column of subplots with arrowhead, bottom to top). We next explored the effects of varying both *k*_3_ and *k*_5_ simultaneously.

**Figure 3:**
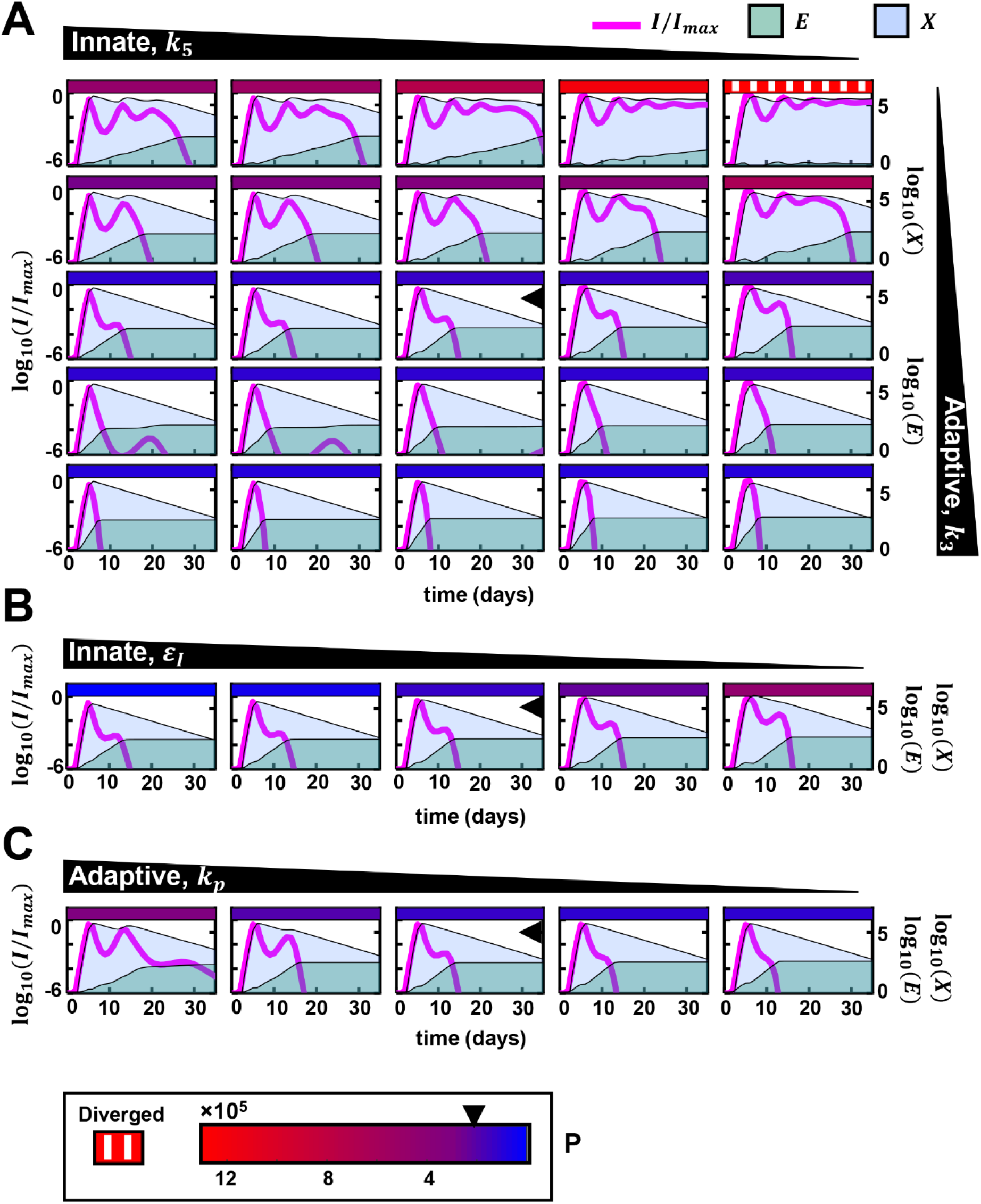
Variations in innate and CD8 T-cell responses capture disease heterogeneity. (A) Effect of simultaneous variation of parameters determining the strengths of innate and CD8 T-cell responses. The black, annotated triangles at the top and right indicate the nature and the direction of the variation of the indicated parameters. For instance, *k*_5_ is decreased from left to right. Individual subplots show the dynamics of infected cells normalized by carrying capacity, cytokine mediated innate immune response and effector CD8 T-cell mediated adaptive immune response. The legends are provided at the top-right. The left Y-axis shows the normalized infected cell dynamics. The right Y-axis shows the other two species, i.e., cytokine-mediated innate immune response and effector CD8 T-cell-mediated adaptive immune response. The rectangular, colored patch at the top of each subplot represents the extent of immunopathology. The range of immunopathology is given by the color scale at the bottom of the figure (below 3C). At the left of the color scale, a separate legend denotes the texture used for depicting diverged immunopathology. The arrowhead on the scale indicates the immunopathology for the population parameters. The central subplot, which also contains an arrowhead, charts the simulation using the population parameters (supplementary table S6). (B, C) Similar calculations corresponding to variations in other parameters associated with the innate (B) and CD8 T-cell (C) responses. Plots with the population parameters are marked with the arrowhead. The colored patches should be interpreted using the color scale provided at the bottom.

When *k*_3_ was high, *i*.*e*., the response of CD8 T-cells was strong, irrespective of the innate immune response, the infection was cleared within ten days (figure 3A, subplots on bottom-left and bottom-right). Associated immunopathology was nominal. These predictions were akin to asymptomatic and mild infection scenarios. An early and robust effector T-cell response has been associated with milder infections (12, 16, 46). Here, in some cases with high *k*_5_, a blip of the viral load was observed after an initial phase of clearance. When *k*_5_ was decreased, marking a weaker innate response, the peak viral load rose and immunopathology moderately increased. This was also observed when we decreased *ε*_*I*_, which lowered the efficacy with which the innate immune response inhibits the spread of the infection (figure 3B). The latter trends associated with high *k*_3_ and low *k*_5_ have parallels to infected patients with robust CD8 T-cell responses but impaired innate responses, such as those harboring mutations in the genes associated with the activation of the antiviral resistance in host cells (47). Clearance was achieved in such cases due to the robust CD8 T-cell response.

When *k*_3_ was low and *k*_5_ was high (figure 3A, four subplots on top-left), the infection was prolonged. However, the immunopathology was lower than when both *k*_3_ and *k*_5_ were high. The efficient antiviral innate response controlled the initial peak of the infection. However, the slow proliferation of the effector cells delayed clearance. This scenario had parallels to the reported cases of prolonged RT-PCR positivity of viral loads (22–24). Restrained CD8 T-cell differentiation was associated with such cases (22). Delayed clearance was also realized when the parameter *k*_*p*_ was increased, which increased the antigen level required for significant effector T-cell proliferation (figure 3C). These predictions were consistent with observations of defects in T-cell proliferation delaying the clearance of infection (22).

When both *k*_3_ and *k*_5_ were low (figure 3A, four subplots on top-right), severe immunopathology along with prolonged infection with high viral load and high cytokine levels was predicted. When they were the lowest, clearance was not achieved in our simulations. Although, clearance of the infection is the predominant outcome associated with a wide range of parameter values (figure 3), our dynamical systems analysis revealed that in certain parameter regimes clearance may not result (supplementary text section D, supplementary figures S6, S7). Instead, escape from immune protection with a high level of infected cells and cytokines together with a high degree of CD8 T-cell exhaustion may occur. Such runaway trajectories were associated with high immunopathology in our model (figure 3A, top right corner, supplementary figure S3B, S3C, top right corner) and were predicted to be terminated by fatality. These trends in the model mirrored clinical features of severe COVID-19 (45), which include consistently very high viral loads, heightened proinflammatory cytokines and interferons (39, 45, 48), attenuated proliferation (13) and increased exhaustion of T-cells (13, 14, 17). The predictions also include cases where in the late phase of the infection, although the viral load in sputum shows a decline (49), and that in nasopharyngeal swab becomes undetectable (33, 39, 49), mortality results due to intolerable immunopathology.

Note that the initial pool of CD8 T-cells, *E*_0_, was important in determining outcomes (supplementary text section D, supplementary figure S7), with a large pool leading to rapid clearance, in agreement with observations of such clearance facilitated by cross-reactive effector T cells (12, 50). The outcomes were less sensitive to the viral inoculum size (supplementary text, section E, supplementary figure S8), i.e., *I*_0_, consistent with studies on macaques where different inoculum sizes led to comparable disease outcomes (51).

### Model can recapitulate clinical data of varying disease severity across patients

In a recent study, patients were stratified by disease outcome and measurements of longitudinal viral load from their saliva were fit using cubic splines, yielding ribbons of confidence intervals on viral loads for each category (39). We digitized these ribbons and tested our model predictions against them (figure 4A-D, grey patches). The study reported data from symptom onset. We therefore added an estimated length of the prodromal period to the timepoints in order to compare our model predictions. We set this length to 4.8 d from the German transmission chain data (37, 38) (supplementary table S2), which is also consistent with other reported estimates (33). We estimated the viral load, *V*, from our model predictions using the pseudo-steady state approximation, *V* ≈ *pI*/*c*, where *p* is the per capita rate of viral production from infected cells and *c* is the per capita rate of viral clearance. We set *p* and *c* to values estimated previously (52) (supplementary text section A).

**Figure 4:**
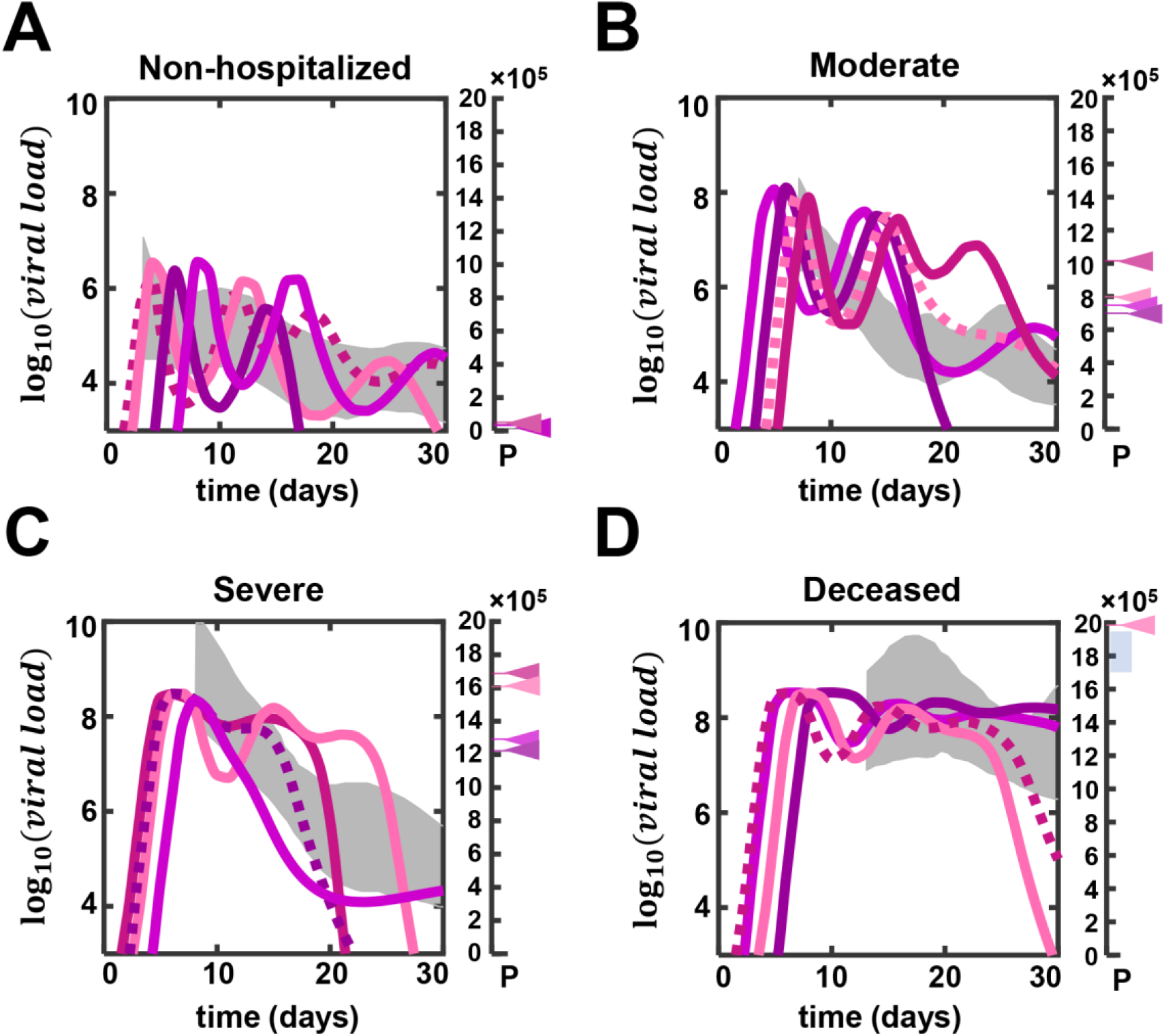
Model recapitulates viral load data from patients with different severities of infection. (A) The grey patch, as provided in Silva et al. (39), represents the confidence interval of a cubic spline fitted to the viral load data from non-hospitalized individuals. Curves depict simulated viral load trajectories in individuals representative of those in the patch. The scale on the right represents immunopathology. The immunopathology associated with the simulated trajectories are marked on the scale using arrowheads having the same colors as the profiles. (B-D) Simulations recapitulating the viral load trajectories from moderate, severe and deceased patients, respectively. Immunopathology of deceased individuals not shown were higher than the upper limit of the scale shown here. The blue bar on the scale in (D) represents the range of immunopathology beyond which fatality is the likely clinical outcome. The parameters used for each trajectory are in supplementary tables S9-S12. Those associated with the profiles with dashed lines were used for sensitivity analyses (supplementary figures S9-S12).

The ribbons of data are not amenable to fitting. We therefore varied parameters associated with the innate and CD8 T-cell responses in our model to achieve dynamical profiles of viral load resembling the ribbons. For the purpose of these simulations we ignored both the potential adverse and favorable effects of antibodies. The ribbon for the non-hospitalized patients was associated with low viral loads (figure 4A). The peak viral load was approximately 10^6^ copies of viral RNA. This relatively low peak viral load could be captured by our model when the strength of the innate response was increased, which we achieved by increasing *k*_5_ and/or *ε*_*I*_ (supplementary table S9). The duration of the infection was dependent on the CD8 T-cell response. Strong CD8 T-cell stimulation, achieved with a low value of *k*_*p*_, led to rapid clearance, whereas weaker stimulation, corresponding to a higher *k*_*p*_, allowed the infection to remain for an extended period. We calculated the immunopathology associated with these simulations and found it to be low (figure 4A, scale on the right; also see below).

The ribbon for patients eliciting moderate symptoms had a relatively higher viral load at the peak, reaching approximately 10^8^ copies of viral RNA (figure 4B). Parameters sufficiently close to the population estimates above allowed us to capture the dynamics for these patients (supplementary table S10). The associated immunopathology was considerably higher than the non-hospitalized patients. For severely infected patients, the viral load peak was above 10^8^, reaching as high as 10^10^ copies (figure 4C). We achieved this high peak viral load by lowering the strength of the innate response (decreasing *k*_5_ and/or *ε*_*I*_; see supplementary table S11). The delayed clearance could be recapitulated by lowering CD8 T-cell stimulation (increasing *k*_*p*_ and/or decreasing *k*_3_). The immunopathology was higher than those calculated to capture the viral load dynamics in moderate patients (compare the scales in figure 4B and 4C). Lastly, for the deceased individuals, the peak viral load was similar to the severe patients. However, the downward trend in the viral load after the peak seen with the severely infected patients was no longer apparent (figure 4D). The viral load remained around 10^8^ RNA copies till day 30 post-exposure. A much weaker innate response (low *ε*_*I*_) and a weaker CD8 T-cell response (high *k*_*p*_) could generate matching profiles (supplementary table S12). The immunopathology for the deceased patients was consistently higher than the severe patients, indicating that there might be an upper limit to the extent of immunopathological tissue damage that lay somewhere between our estimates for severe and deceased patients, and crossing which mortality would almost certainly result.

Our model thus recapitulated the trends in the viral load seen in patients with different severity of infection. Furthermore, the model indicated that there should be a narrow range of immunopathology, which acts as a threshold to determine the fatal outcomes in COVID-19 (figure 4D, scale on the right).

To assess whether variations in other model parameters could also achieve the above trends, we performed a global sensitivity analysis of our parameters using one representative parameter combination from each disease outcome category as reference (figure 4A-D, dashed curves; supplementary figures S9-S12; supplementary tables S9-S12). Specifically, we calculated how sensitive our measure of immunopathology was to the parameters. We found that immunopathology was most sensitive to *ε*_*I*_ and *k*_6_ for non-hospitalized and moderately symptomatic patients (supplementary figures S9, S10). For severely infected and deceased patients, *I*_*max*_, *k*_3_ and *ε*_*I*_ emerged as the important parameters (supplementary figures S11, S12). These results reinforce our expectations above. In mild infections, the innate immune response is strong and any variation in its strength would have the most influence on disease severity. In more severe infections, the innate and adaptive responses are both involved and the severity is therefore sensitive to variations in the strengths of both.

Model predictions thus successfully recapitulated the heterogeneous outcomes and the associated dynamical patterns of SARS-CoV-2 infection.

## Discussion

The extreme heterogeneity in the outcomes of SARS-CoV-2 infection across infected individuals has been puzzling. Here, using mathematical modeling and analysis of patient data, we predict that the heterogeneity arises from the variations in the strength and the timing of the innate and the CD8 T-cell responses across individuals. When both the innate and the CD8 T-cell arms are strong, asymptomatic or mild infections result. When the CD8 T-cell arm is strong, clearance of the infection results. If the innate arm is weak, the peak viral load can be large, resulting in higher immunopathology and moderate symptoms. When the CD8 T-cell response is strong but delayed, a predator-prey type interaction between the innate arm and the virus results, causing multiple peaks in viral load. These oscillations end when the CD8 T-cell response is mounted, and clearance ensues. When the CD8 T-cell response is weak, but the innate arm is strong, prolonged infection can result before clearance. When both the arms are weak, severe infection including mortality follows. These predictions offer a conceptual understanding of the heterogeneous outcomes of SARS-CoV-2 infection. They also offer a synthesis of the numerous independent and seemingly disconnected clinical observations associated with the outcomes and present a framework that may help tune interventions.

In the last year, several mathematical models of within-host SAR-CoV-2 dynamics have been developed and have offered valuable insights (53). For instance, they have helped estimate the within-host basic reproductive ratio (33, 34, 52) and assess the effects of drugs and vaccines (26, 27, 44, 54–57). Attempts have also been made to capture the role of the immune system in disease progression and outcome (44, 55, 57–61). Available models, however, have either not been shown to fit longitudinal patient data or have failed to describe the entire range of outcomes realized. To our knowledge, ours is the first study to describe the outcomes realized comprehensively using a mathematical model that is consistent with patient data.

Our model predictions help better understand known demographic correlates of disease severity and mortality, such as gender, age and co-morbidities. In all these cases, as our predictions indicate, more severe infections are associated with weaker CD8 T-cell responses and/or unregulated innate immune responses. Male patients trigger higher levels of peripheral cytokine expression and elicit weaker CD8 T-cell responses than female patients (62), resulting in more frequent severity and mortality in males (43). The increased mortality in the elderly is caused by immunosenescence, which is associated with decreased proliferative capacity of lymphocytes and impaired functionality of innate immune cells (63). Increased mortality is also associated with co-morbidities, such as type-2 diabetes (64), where uncontrolled production of proinflammatory cytokines and inappropriate recruitment of lymphocytes is observed (65).

Factors in addition to the above could contribute to variations in the innate and the CD8 T-cell responses across individuals. For instance, certain mutations, reported in a subset of severe COVID-19 patients, may preclude a potent interferon response (47). A section of severely infected patients is reported to harbor neutralizing autoantibodies against interferons (29, 66). Overzealous production of antibodies against SARS-CoV-2 might inhibit the pathway for interferon-mediated induction of antiviral genes (67). Further, in vitro studies suggest that different SARS-CoV-2 proteins can inhibit the TBK1-IRF3 pathway or the JAK/STAT pathway at several signaling nodes, adversely affecting interferon production and/or signaling (68). Variability in the CD8 T-cell response may come from different precursor populations, due for instance to prior exposure to circulating human coronaviruses (50). Patients pre-exposed to other coronaviruses or rhinoviruses harbor populations of effector T-cells that might cross-react with SARS-CoV-2 antigens and contribute to the early clearance of the infection (50, 69). Population-level variations in effector cell frequencies (70) and inter-individual heterogeneity in lymphocytic gene expression patterns (71) may also contribute to the variability in the CD8 T-cell response.

CD8 T-cell exhaustion has been proposed as an evolutionary design to prevent mortality due to immunopathology (30, 72). By preventing extensive tissue damage due to CD8 T-cell killing of infected cells, exhaustion can avert mortality. The price of reduced CD8 T-cell efficiency is often persistent infection, as seen with HIV and hepatitis C (30). With severe SARS-CoV-2 infection, although extensive CD8 T-cell exhaustion is seen, it appears inadequate to prevent mortality; immunopathology caused by proinflammatory cytokines dominates. Potent activation of the NF-κB pathway by components of the SARS-CoV-2 virion may trigger the production of detrimental proinflammatory cytokines (73, 74). Heightened interferon expression in the lung (5, 18, 19, 75, 76) impairs cell proliferation, impeding tissue repair after proinflammatory cytokine-mediated immunopathology (77). Moreover, interferons may synergize with proinflammatory cytokines to fuel immunopathology by triggering cell death pathways (78, 79). (Note that interferons may be subdued in peripheral circulation (80), but that appears to be uncorrelated with their expression in the respiratory tract in COVID-19 (5).) In contrast, immunopathology due to CD8 T-cells appears minimal. CD8 T-cells infiltrate the alveolar tissues of COVID-19 patients (76) and can kill infected cells. At the peak of the infection, 10^4^-10^6^ cells are estimated to be infected out of the 10^11^ estimated target cells in the respiratory tract (81). Thus, direct CD8 T-cell killing of infected cells would affect a small fraction of cells in the respiratory tract. This may also explain why viral persistence has not been observed with SARS-CoV-2 infection: Inducing CD8 T-cell exhaustion can only minimally affect immunopathology dominated by cytokines. We speculate that the absence of persistence may be a general feature of those viral infections where immunopathology is predominantly cytokine-mediated. Indeed, hypercytokinemia has been associated with the fatal outcomes following influenza A (H5N1) infection (82).

A strategy of great interest today for reinvigorating exhausted CD8 T-cells is the use of immune checkpoint inhibitors (83). The inhibitors are approved for use in certain cancers. Because of their promise, five clinical trials are underway for testing their efficacy in treating severe COVID-19, of which one (NCT04333914) is on cancer patients, and the remaining (NCT04413838, NCT04343144, NCT04356508, and NCT04268537) are on non-cancer patients infected by SARS-CoV-2 (84). A major risk of checkpoint inhibitor therapy is increased immunopathology due to a heightened CD8 T-cell response. Based on our model predictions and arguments above, we speculate that with COVID-19, the risk of increased immunopathology from immune checkpoint inhibitor therapy is likely to be minimal, given the predominance of cytokine-mediated pathology. Indeed, a retrospective analysis of melanoma patients showed that checkpoint inhibitor therapy did not increase the risk of mortality due to COVID-19 (85). Rather, the beneficial effects of an improved CD8 T-cell response may outweigh any minimal enhancement in immunopathology.

Our model could be applied to understand the implications of other interventions (86) and of emerging viral mutants (87) on disease outcomes. Given the mechanisms of action of available drugs and drug candidates (86), their effects on typical individuals in the mild, moderate and severe infection categories could be predicted using the corresponding nominal parameter estimates we identified for the respective categories. Several recently identified circulating mutants are known to be more infectious than the original SARS-CoV-2 strain and to escape immune responses (88). These characteristics could be incorporated in our model readily by suitably increasing the infectivity and/or decreasing the strength of the immune response, to predict how emerging strains could alter the overall severity of the infection. We recognize that to estimate the effects of such variations at the population level, knowledge of how the parameter values in our model, particularly those defining the innate and CD8 T-cell responses, are distributed across individuals in a population would be required. With hepatitis C virus infection, for instance, the distribution of the strength of interferon responsiveness across individuals quantitatively predicted the fraction of individuals that spontaneously cleared the infection (89, 90) and together with the distribution of the CD8 T-cell response captured the success of interferon-based and other therapies (89–91). Such predictions with SARS-CoV-2, once parameter distributions become available, may help refine clinical and epidemiological projections of healthcare requirements.

Our study has limitations. First, we neglected the role that cytokines play in the expansion of CD8 T-cells (92) because fits of our model incorporating such an effect to the available data were poor (supplementary text section B). Perhaps, a larger patient cohort may improve the fits and allow incorporating the latter effect. Second, our model did not incorporate any negative effect of immunopathology on the immune response; for instance, lymphopenia (15, 93), which is generally thought to be caused by immunopathology, could compromise the immune response. Third, we employed a simplified model of CD8 T-cell exhaustion, following earlier studies (30–32), which allows exhaustion to be reversed fully upon lowering antigen levels. Recent studies have demonstrated that exhaustion is reversible only in a subset of exhausted cells (83). Notwithstanding, we expect our key inferences on the roles of the innate and the CD8 T-cell responses in determining the heterogeneous outcomes of SARS-CoV-2 infection to hold.

## Methods

### Study design

We constructed a mathematical model of within-host SARS-CoV-2 infection using ordinary differential equations. Next, we utilized a nonlinear mixed-effects approach to fit the model to an available clinical dataset and estimated model parameters (supplementary text, section A, B). The model was then utilized for exploring the effects of parameter variations (figure 3), recapitulating the viral load trajectories in patients stratified by disease severity (figure 4), and for sensitivity analysis (supplementary figures S9-S12).

### Model construction

The equations of our model are provided in the results section. The other models which were fit to the data are described in the supplementary text (supplementary text, section B).

### Parameter estimation and model selection

Published data from Wölfel et al.(37) was digitized by a custom script in the MATLAB (version R2020a) image analysis toolbox (www.mathworks.com). This dataset was further used for fitting different models using the stochastic approximation expectation maximization (SAEM) algorithm available in Monolix 2020R1 (www.lixoft.com) (supplementary text, section A). The Akaike information index (AIC) was calculated within the Monolix environment. The model with the lowest AIC was selected for further mathematical analysis (supplementary text, section B).

### Fixed points and linear stability analysis

For the steady-state analysis, estimated parameter values were utilized. MATLAB (version R2020a) was used to estimate the fixed points of the system and to determine the nature of their stability. Individual fixed points and their corresponding Jacobian matrices were estimated using the Symbolic Math Toolbox (www.mathworks.com). Calculation of the eigenvalues and eigenvectors for individual fixed points yielded the nature of their stability and facilitated determination of the phase portraits (supplementary text, section D, supplementary figures S6, S7).

### Recapitulating patient viral load data stratified by disease severity

The published data from Silva et al. (39), was digitized using a custom MATLAB code, using functions from the image analysis toolbox. The parameters were manually varied and the quality of the fits determined by visual inspection of the simulation profile and the confidence interval ribbons.

### Sensitivity analysis

We executed variance based global sensitivity analysis (VBGSA) on the models; the details of the algorithm have been described elsewhere (94). We simultaneously varied the parameters up to 5% above and below the population parameters in Monte Carlo simulations and calculated total effect indices for the parameters (supplementary figures S9-S12).

## Data Availability

All data used in the study are provided in the main text and supplementary material.

## Acknowledgments

We thank Pranesh Padmanabhan for insightful comments and Rajat Desikan for help with the Monolix platform. BC is supported by the C. V. Raman postdoctoral fellowship at the Indian Institute of Science.

## Author contributions

Conceptualization: BC, HSS, NMD

Methodology: BC, HSS

Investigation: BC, HSS

Visualization: BC, HSS

Supervision: NMD

Writing—original draft: BC, HSS

Writing—review & editing: BC, HSS, NMD

## Data and materials availability

All data used in the study are provided in the main text and supplementary material. Codes are available upon request.

