## Supplementary Materials for "The relative strength and timing of innate immune and CD8 T-cell responses underlie the heterogeneous outcomes of SARS-CoV-2 infection"

[†]These authors contributed equally

This file includes:

A. A detailed description of the fitting procedure

Table S1: Events of exposure of the patients

Table S2: Time of symptom onset in the patients

Table S3: Dates of first measurements of viral loads in patients

Table S4: Comparison of the fixed effects and their standard errors for the model with and without $I_{max}$

Table S5: Fitting the model without $I_{max}$ to different initial conditions of $i$

Table S6: Estimated population parameters

Table S7: Estimated individual parameters

B. A detailed description of model selection

Table S8: Information indices of candidate models

Figure S1: Infection trajectory when $E_0 = 0$

Figure S2: The individual fits are not affected when simulated with estimated $k_4$ and $k_e$

Figure S3: The schema of immunopathology calculation

Figure S4: Change in the dynamics of innate response and that of infection with change in $k_5$

C. Sensitivity of Immunopathology towards $\alpha, \beta, \gamma$

Figure S5: Sensitivity estimates of perturbations in the parameters associated with immunopathology

D. A detailed description of the stability analysis

Figure S6: Flow chart of categories in phase portraits

Figure S7: Monostability and bistability, sample trajectories and associated immunopathology

E. Effect of varying viral inoculum size

Figure S8: Similar outputs with varying starting viral inoculum

Table S9-S12: The parameters generating the simulations recapitulating viral loads

Figure S9-S12: Global sensitivity analysis of the model parameters recapitulating viral loads in patients of different severity

### A. Detailed description of the fitting procedure

#### A.1. Data collection and curation

We collected the dates of exposures of all patients from the report by Böhmer et al. (1) (table S1). Some patients had multiple events of exposure on consecutive days. We selected the first day of exposure as day 0, assuming that the viral transmission occurred on the first exposure. Further, we collected the dates of symptom onset for all patients and calculated the days post-infection associated with those dates (table S2). Next, we digitized the viral load data of these patients from the figure 2 of the report by Wölfel et al.(2). In the latter figure, viral load is plotted versus days post symptom onset. Combining the latter with duration for symptom onset, we deduced the dates of the first viral load measurements, or diagnosis (table S3), allowing us to determine viral load as a function of the time from the first exposure, which is presented in our figure 2. We thus collated data from day 0, i.e., the date of exposure, to day 15 into the infection. The earliest diagnostic data was available for patient 3, on day 4 post exposure. The latest diagnostic data was provided for patient 8, on day 12 post exposure. All dates provided in the tables are for the year 2020.

**Table S1: Events of exposure of the patients**

| Patient ID | First exposure (day 0) | Repeated exposure (if any) (day #) | Reference |
|---|---|---|---|
| 1 | 20 Jan | 0,1 | Page 923, 3rd paragraph, "patient 1 was…on the same day." Böhmer et al. (1) |
| 2 | 20 Jan | 0,1,2 | We assume the earliest possible date of transmission. No direct contact traced with patient 0. Viral genomic sequencing established the source. Table 1. Böhmer et al., (1). Air borne spread of the virus might explain such observation (3). |
| 3 | 24 Jan | - | Figure; Böhmer et al. (1) |
| 4 | 20 Jan | 0,1,2 | Figure; Böhmer et al. (1) |
| 7 | 24 Jan | - | Figure; Böhmer et al. (1) |
| 8 | 22 Jan | 0,1,2 | Page 923, 7th paragraph, "patient 7 met…on the day of testing". We assume that the transmission occurred while patient 5 was in prodromal phase. Böhmer et al., (1) |
| 10 | 28 Jan | - | Figure; Böhmer et al. (1) |
| 14 | 28 Jan | 0,1,2,3 | Figure; Böhmer et al. (1) |

**Table S2: Time of symptom onset in the patients**

| Patient ID | Symptom onset (day post exposure) | Reference |
|---|---|---|
| 1 | 23 Jan (day 3) | Page 923, 3rd paragraph, "patient 1 was…on the same day." Böhmer et al. (1) |
| 2 | 25 Jan (day 5) | Figure; Böhmer et al. (1) |
| 3 | 25 Jan (day 1) | Figure; Böhmer et al. (1) |
| 4 | 24 Jan (day 4) | Figure; Böhmer et al. (1) |
| 7 | 28 Jan (day 4) | Figure; Böhmer et al. (1) |
| 8 | 28 Jan (day 6) | Figure; Böhmer et al. (1) |
| 10 | 30 Jan (day 2) | Figure; Böhmer et al. (1) |
| 14 | 03 Feb (day 6) | Figure; Böhmer et al. (1) |

**Table S3: Dates of first measurements of viral loads in patients**

| Patient ID | First test (day post exposure) | Reference |
|---|---|---|
| 1 | 27 Jan (day 7) | Page 923, 3rd paragraph, "patient 1 was…on the same day." Böhmer et al. (1) |
| 2 | 28 Jan (day 8) | Calculated from figure 2, Wölfel et al. (2) |
| 3 | 28 Jan (day 4) | Calculated from figure 2, Wölfel et al. (2) |
| 4 | 28 Jan (day 8) | Calculated from figure 2, Wölfel et al. (2) |
| 7 | 01 Feb (day 8) | Calculated from figure 2, Wölfel et al. (2) |
| 8 | 03 Feb (day 12) | Calculated from figure 2, Wölfel et al. (2) |
| 10 | 03 Feb (day 6) | Calculated from figure 2, Wölfel et al. (2) |
| 14 | 05 Feb (day 8) | Calculated from figure 2, Wölfel et al. (2) |

### A.2. Description of the fitted model

We fit the following model equations to the data:

$$\frac{dI}{dt} = [k_1(1 - \varepsilon_I X)I\left(1 - \frac{I}{I_{max}}\right) - k_2 IE]H(t - \tau)$$

$$\frac{dE}{dt} = [k_3\left(\frac{1}{k_p + I}\right)IE - k_4\left(\frac{1}{k_e + I}\right)IE]H(t - \tau)$$

$$\frac{dX}{dt} = [k_5 I - k_6 X]H(t - \tau)$$

The descriptions of the parameters are provided in the results section of the main text. For the purpose of fitting, we introduced the Heaviside function, $H(t - \tau)$, which equals 1 when $t > \tau$ and 0 otherwise, to account for the delay in viral replication post exposure, $\tau$. Visual inspection of the dataset indicated that at least for some patients, the viral load did not start rising immediately after exposure. The dynamical events of the infection were thus initiated after the duration $\tau$, which we estimated from the fits. Further, as elaborated in the results section, we fixed $k_4 = 0$. We further assumed that $k_5 = 1$, to reduce the number of parameters without loss of generality. As detailed in the results section, we assumed $I_{max} = 10^6$, for all patients. We tested the sensitivity to the latter assumption below.

### A.3. Examining the dependence of the fits on $I_{max}$

The above equations could be readily simplified to the following:

$$\frac{di}{dt} = [k_1(1 - \varepsilon_I' x)i(1 - i) - k_2 iE]H(t - \tau)$$

$$\frac{dE}{dt} = [k_3\left(\frac{1}{k_p' + i}\right)iE - k_4\left(\frac{1}{k_e' + I}\right)]H(t - \tau)$$

$$\frac{dx}{dt} = [k_5 i - k_6 x]H(t - \tau)$$

where $i = I/I_{max}$, $x = X/I_{max}$, $k_p' = k_p/I_{max}$, and $\varepsilon_I' = \varepsilon_I I_{max}$. The explicit dependence of the dynamics on $I_{max}$ thus vanishes, except through the initial condition for $i$. We fit the data using the latter equations and found that the best-fit parameter estimates were indistinguishable from those obtained earlier (table S4). Further, we fit this simplified model with different initial values of $i$ and found no significant differences in the parameters estimated (table S5). Our model and fits were thus not sensitive to the choice of $I_{max}$.

**Table S4: Comparison of parameter estimates for the models with and without explicit $I_{max}$**

| Parameters | Model with ***I*** | | The simplified model with ***i*** | |
|---|---|---|---|---|
| | Fixed effect | S.E. | Fixed effect | S.E. |
| $k_1$ | 4.6 | 1.71 | 4.66 | 1.39 |
| $-\log_{10} k_2$ | 1.65 | 0.25 | 1.66 | 0.75 |
| $k_3$ | 0.64 | 0.041 | 0.68 | 0.28 |

| | | | | |
|---|---|---|---|---|
| $k_6$ | 0.24 | 0.039 | 0.26 | 0.031 |
| $\log_{10} E_0$ | 0.1 | 0.00053 | 0.1 | 0.005 |
| $-\log_{10} \varepsilon_I$ | 5.5 | 0.68 | 5.4 | 0.48 |
| $\log_{10} k_p$ | 2.04 | 0.38 | 2.02 | 0.32 |
| $\tau$ | 1.9 | 1.19 | 2.3 | 1.36 |

**Table S5: Sensitivity of the fit parameters to different initial values of $\boldsymbol{i}$**

| Parameters | $\boldsymbol{i_0}$ **= 10$^{-7}$** | | $\boldsymbol{i_0}$**= 10$^{-6}$** | | $\boldsymbol{i_0}$ **= 10$^{-5}$** | |
|---|---|---|---|---|---|---|
| | Fixed effect | S.E. | Fixed effect | S.E. | Fixed effect | S.E. |
| $k_1$ | 4.62 | 1.26 | 4.66 | 1.39 | 4.41 | 1.44 |
| $-\log_{10} k_2$ | 1.57 | 0.44 | 1.66 | 0.75 | 1.65 | 0.27 |
| $k_3$ | 0.64 | 0.11 | 0.68 | 0.28 | 0.67 | 0.1 |
| $k_6$ | 0.25 | 0.032 | 0.26 | 0.031 | 0.26 | 0.036 |
| $\log_{10} E_0$ | 0.1 | 0.022 | 0.1 | 0.005 | 0.1 | 0.021 |
| $-\log_{10} \varepsilon_I$ | 5.48 | 0.54 | 5.4 | 0.48 | 5.34 | 0.52 |
| $\log_{10} k_p$ | 2 | 0.026 | 2.02 | 0.32 | 2.05 | 0.68 |
| $\tau$ | 1.39 | 1.31 | 2.3 | 1.36 | 2.82 | 1.2 |

#### A.4. Fitting using Monolix

To ensure that the fitting captured the basic trends of the viral dynamics, we censored the peaks in the data for each patient. This was for ensuring that the data below the peak received weightage in the fitting. For patient 1 and 2, where a relatively longer viral incubation was evident from visual inspection of the data, we censored the first few days so that the viral load did not rise in these early time points. We used logit distributions for fitting all parameters and supplied biologically relevant ranges for them. The fitted population parameters (table S6) and individual parameters (table S7) were collected from Monolix, and further simulations and analyses were run in MATLAB.

**A.5. Selection of parameters not estimated in the fitting**

In our fitting exercise we ignored the parameters associated with exhaustion and immunopathology. We obtained the latter parameters as follows. We chose $k_4$ from a previously published analysis (4). We then chose $k_e$ such that no effect of exhaustion was observed for the simulations corresponding to the best-fits to the patient data (see figure S2). This ensured internal consistency with our assumption and agreement with observations of minimal pathology in mildly infected patients. The parameters for the immunopathology were either taken from a previously published source (4) or assumed. Varying these parameters ten-fold up and down did not alter our inferences (supplementary text, section C). The following values of the parameters were used in all simulations: $k_4 = 1.5$ day$^{-1}$; $k_e = 7e + 05$ cells; $\alpha = 1e - 03$ cells$^{-1}$day$^{-1}$; $\beta = 5e - 02$ day$^{-1}$; $\gamma = 0.5$ day$^{-1}$; The parameters associated with viral dynamics, as shown in figure 4, were taken by averaging the estimates provided in Wang et al. (5); thus, we set $p = 1.78e + 04$ cells$^{-1}$day$^{-1}$ and $c = 50.34$ day$^{-1}$.

**Table S6: Estimated population parameters**

| Parameter | Value | S.E. |
|---|---|---|
| $k_1$ | 4.6 day$^{-1}$ | 1.71 day$^{-1}$ |
| $\varepsilon_I$ | 3.16E-06 | 2.5E-06 |
| $E_0$ | 1.26 | 0.0016 |
| $k_2$ | 0.0224 day$^{-1}$ | 0.0098 day$^{-1}$ |
| $\tau$ | 1.9 days | 1.19 days |
| $k_3$ | 0.64 cells$^{-1}$day$^{-1}$ | 0.041 cells$^{-1}$day$^{-1}$ |
| $k_p$ | 109.65 cells | 153.4 cells |
| $k_6$ | 0.24 day$^{-1}$ | 0.039 day$^{-1}$ |

**Table S7: Estimated individual parameters.** The units are the same as in table S6.

| Patient No. | $k_1$ | $k_2$ | $k_3$ | $k_6$ | $E_0$ | $\varepsilon_I$ | $k_p$ | $\tau$ |
|---|---|---|---|---|---|---|---|---|
| 1 | 4.55 | 0.0224 | 0.64 | 0.24 | 1.26 | 2.63E-06 | 104.7 | 5.18 |
| 2 | 4.65 | 0.0224 | 0.64 | 0.25 | 1.26 | 3.38E-06 | 416.9 | 6.63 |
| 3 | 4.72 | 0.0229 | 0.64 | 0.24 | 1.26 | 5.01E-05 | 166.0 | 0.78 |
| 4 | 4.53 | 0.0224 | 0.64 | 0.25 | 1.26 | 2.63E-06 | 102.3 | 2.71 |
| 7 | 4.61 | 0.0224 | 0.64 | 0.24 | 1.26 | 3.38E-06 | 2398.8 | 2.66 |
| 8 | 4.7 | 0.0224 | 0.64 | 0.25 | 1.26 | 3.23E-06 | 5623.4 | 1.1 |
| 10 | 4.57 | 0.0224 | 0.64 | 0.25 | 1.26 | 3.02E-06 | 2570.4 | 1.23 |
| 14 | 4.43 | 0.0224 | 0.64 | 0.25 | 1.26 | 2.45E-06 | 100 | 1.15 |

### B. Detailed description of model selection

The following candidate models were fit to the data and Akaike Information Criterion (AIC) were determined (table S8). We present the equations and mention the differences from the model presented in the main text.

#### B.1 Model with saturating innate immune response and its priming of effector response

$$\frac{dI}{dt} = [k_1(1-\varepsilon_I X)I\left(1-\frac{I}{I_{max}}\right) - k_2 IE]H(t-\tau)$$

$$\frac{dE}{dt} = [k_3\left(\frac{1}{k_p+I}\right)(1+\varepsilon_E X)IE - k_4\left(\frac{1}{k_e+I}\right)IE]H(t-\tau)$$

$$\frac{dX}{dt} = [k_5 I\left(1-\frac{I}{k_i}\right) - k_6 X]H(t-\tau)$$

Here, in addition to the terms in the model in the main text, the effect of the innate immune response, $X$, on CD8 T-cell stimulation, represented by the term $\varepsilon_E X$, and a maximum limit to the innate response via the carrying capacity $k_i$ were introduced. All the other parameters and variables are the same as in the main text.

#### B.2. Model with saturating immune response

$$\frac{dI}{dt} = [k_1(1-\varepsilon_I X)I\left(1-\frac{I}{I_{max}}\right) - k_2 IE]H(t-\tau)$$

$$\frac{dE}{dt} = [k_3\left(\frac{1}{k_p+I}\right)IE - k_4\left(\frac{1}{k_e+I}\right)IE]H(t-\tau)$$

$$\frac{dX}{dt} = [k_5 I\left(1-\frac{I}{k_i}\right) - k_6 X]H(t-\tau)$$

Here, the saturating innate immune response but not its effect on priming CD8 T-cells in model B.1 was included.

#### B.3. Model without antiviral activity of innate immune response

$$\frac{dI}{dt} = [k_1 I\left(1-\frac{I}{I_{max}}\right) - k_2 IE]H(t-\tau)$$

$$\frac{dE}{dt} = [k_3\left(\frac{1}{k_p+I}\right)(1+\varepsilon_E X)IE - k_4\left(\frac{1}{k_e+I}\right)IE]H(t-\tau)$$

$$\frac{dX}{dt} = [k_5 I\left(1-\frac{I}{k_i}\right) - k_6 X]H(t-\tau)$$

In this model, we removed the term $(1-\varepsilon_I X)$ representing the innate immune response against the spread of the infection, from the dynamics of $I$ in model B.1.

#### B.4. Model without antiviral activity of CD8 T-cells

$$\frac{dI}{dt} = [k_1(1-\varepsilon_I X)I\left(1-\frac{I}{I_{max}}\right)]H(t-\tau)$$

$$\frac{dE}{dt} = [k_3\left(\frac{1}{k_p+I}\right)(1+\varepsilon_E X)IE - k_4\left(\frac{1}{k_e+I}\right)IE]H(t-\tau)$$

$$\frac{dX}{dt} = [k_5 I\left(1-\frac{I}{k_i}\right) - k_6 X]H(t-\tau)$$

Here, we removed the term $k_2IE$, which represents the CD8 T-cell mediated clearance of the infected cells from model B.1.

**B.5. Main model**

$$\frac{dI}{dt} = [k_1(1-\varepsilon_I X)I\left(1-\frac{I}{I_{max}}\right) - k_2IE]H(t-\tau)$$
$$\frac{dE}{dt} = [k_3\left(\frac{1}{k_p+I}\right)IE - k_4\left(\frac{1}{k_e+I}\right)IE]H(t-\tau)$$
$$\frac{dX}{dt} = [k_5I - k_6X]H(t-\tau)$$

This is the model in the main text.

**B.6 Model with innate immune response priming the effector response**

$$\frac{dI}{dt} = [k_1(1-\varepsilon_I X)I\left(1-\frac{I}{I_{max}}\right) - k_2IE]H(t-\tau)$$
$$\frac{dE}{dt} = [k_3\left(\frac{1}{k_p+I}\right)(1+\varepsilon_E X)IE - k_4\left(\frac{1}{k_e+I}\right)IE]H(t-\tau)$$
$$\frac{dX}{dt} = [k_5I - k_6X]H(t-\tau)$$

Here, the effect of the innate immune response on priming CD8 T-cells but not the saturating limit to the innate immune response in model B.1 was included.

We fit the above models to the datapoints shown in figure 2 of the main text (tables S1, S3) following the same procedure as in figure 2 and estimated the AIC. (In particular, we fixed $k_4$ = 0 and $k_5$ = 1 in all cases.)

**Table S8: Values of the Akaike information criterion (AIC) for the candidate models**

| **Model** | **AIC** |
|---|---|
| Model without antiviral activity of innate immune response (B.3) | 274.71 |
| Model without antiviral activity of CD8 T-cells (B.4) | 260.91 |
| Model with innate immune response priming the effector response (B.6) | 253.1 |
| Model with saturating immune response (B.2) | 243.91 |
| Model with saturating innate immune response and its priming of effector response (B.1) | 241.52 |
| Main model (B.5) | 234.72 |

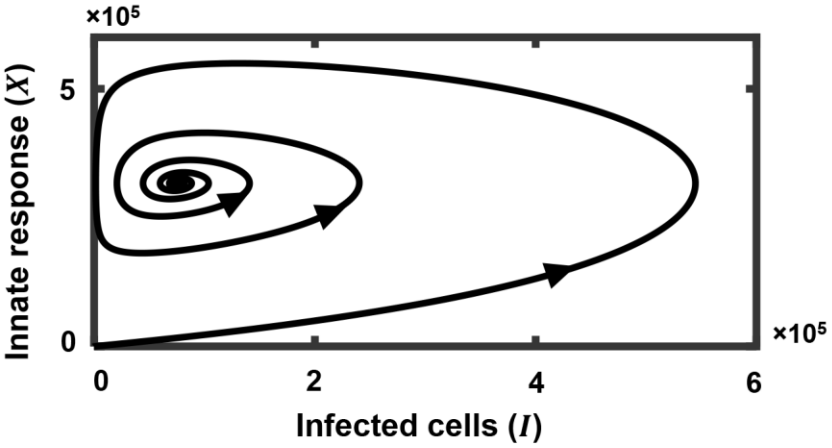


**Figure S1: Infection dynamics without CD8 T-cells.** We solved model equations setting the initial CD8 T-cell pool to zero ($E_0 = 0$), which precluded the emergence of a CD8 T-cell response in our model. The resulting trajectory shows the predator-prey type oscillations, ultimately reaching one of the persistence states (fixed points 2 or 3; see section D).

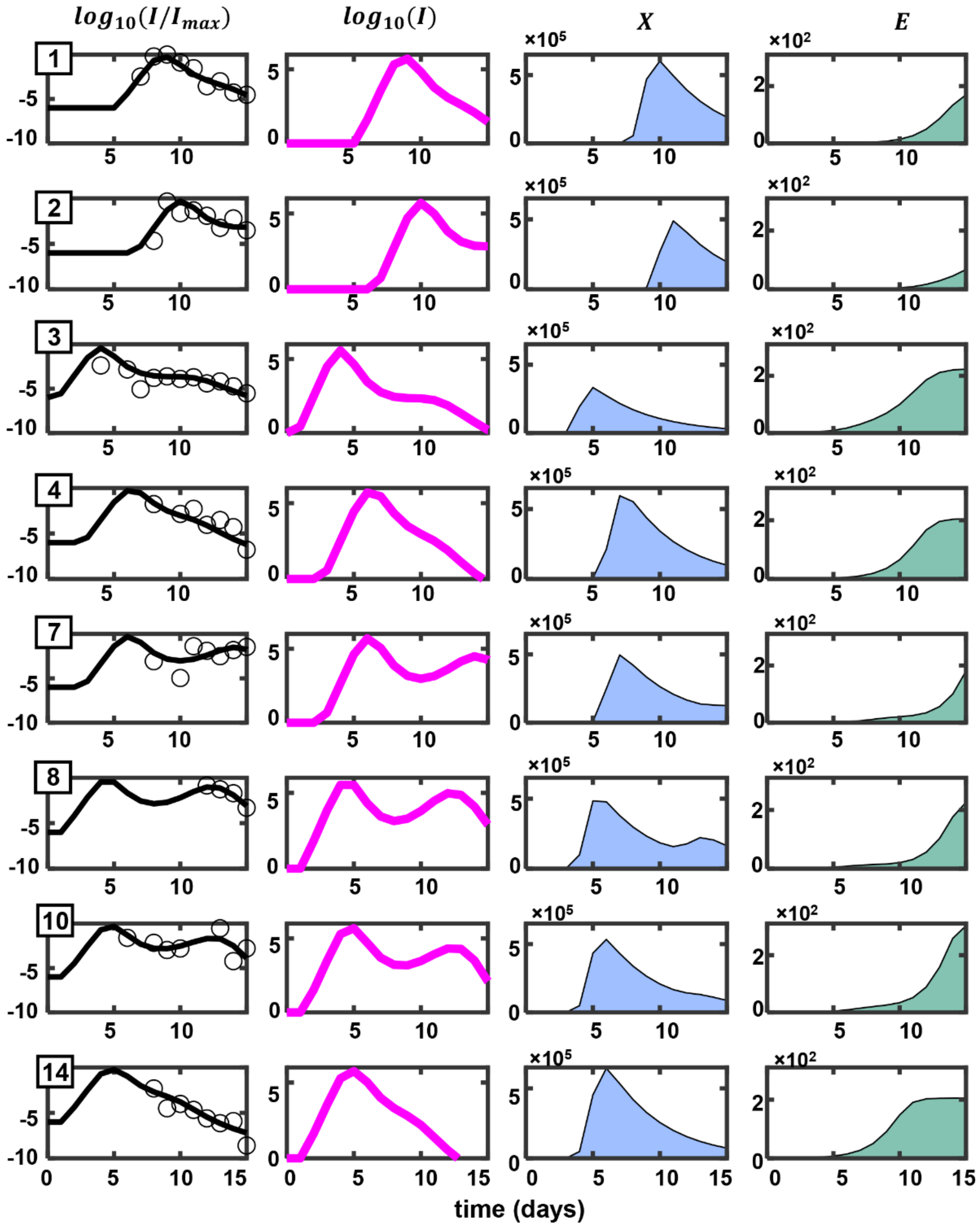


**Figure S2**: **The fits are not affected upon reintroducing CD8 T cell exhaustion.** We recalculated the dynamics following reintroduction of the CD8 T-cell exhaustion term using best-fit parameters for each patient and the chosen values of $k_4$ and $k_e$ (section A.5). The predictions are indistinguishable from those in figure 2. The panels and the quantities depicted are all identical to those in figure 2.

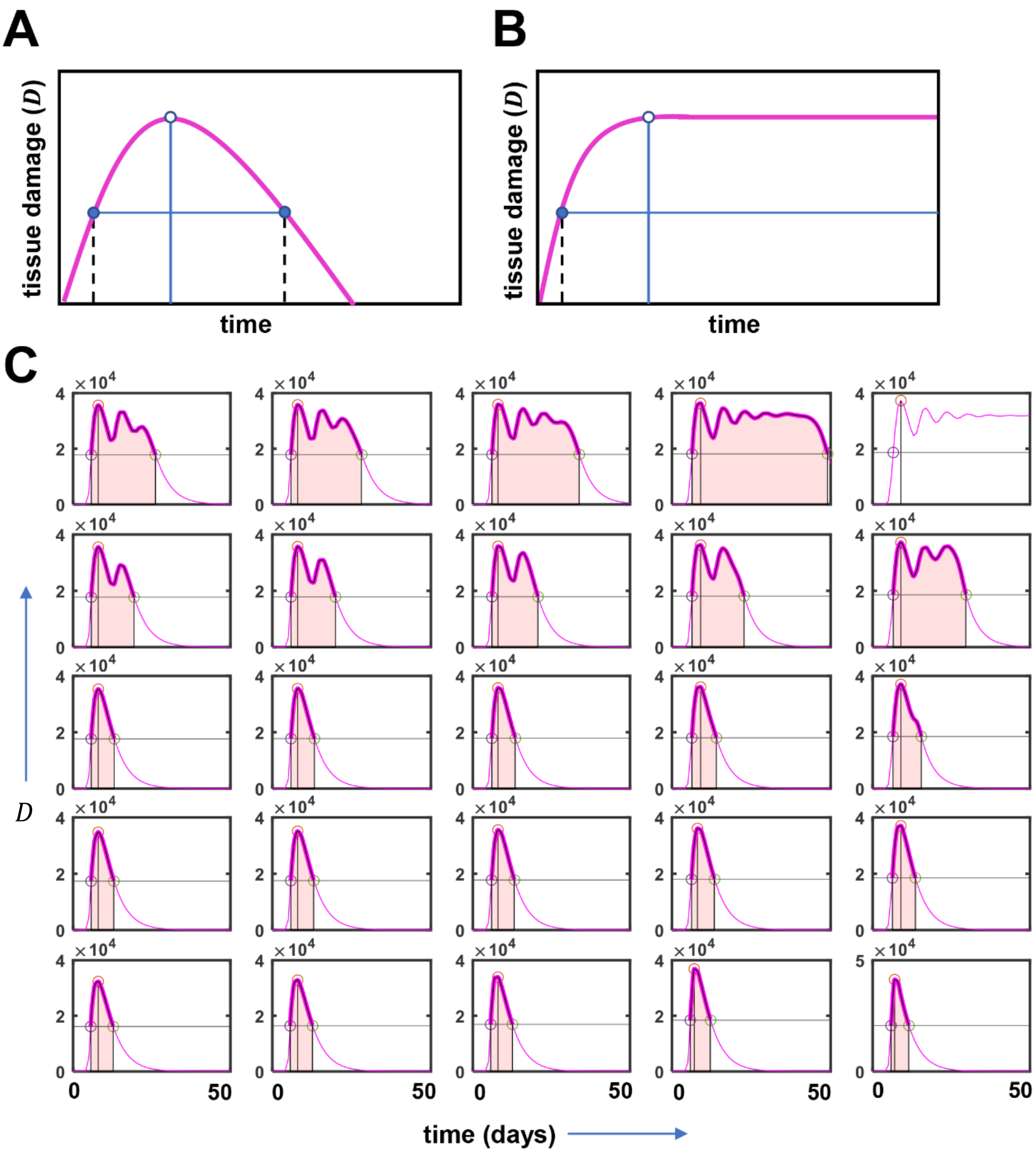


**Figure S3: The schema for the calculation of immunopathology.** (A) The first peak of the tissue damage, $D$ (magenta curve), was detected. A horizontal line was drawn at the half-maximal level of the peak of tissue damage. The two intercepts of the magenta curve with the horizontal line were identified. The area under the magenta curve (AUC) was calculated within the half-maximal intercepts. (B) For cases where the tissue damage saturates to a positive value, we set the immunopathology as 'diverged'. (C) Immunopathology ($P$) calculations for figure 3A in the main text. The area shaded light pink in each panel represents the calculated AUC.

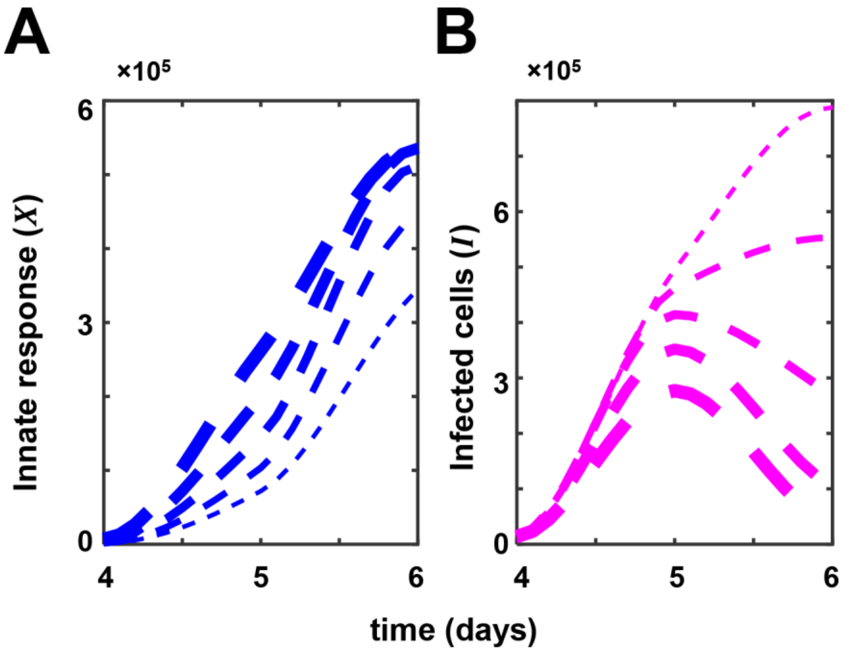


**Figure S4: Sensitivity of the dynamics of innate cytokine response to $k_5$.** The width of the curves is proportional to the strength of $k_5$. (A) and (B) represent the dynamics of $X$ and $I$, respectively. Curves of the same widths represent outputs from the same model.

### C. Effect of perturbations in parameters associated with immunopathology

To test if our results are sensitive to variations in $\alpha, \beta$ or $\gamma$, we estimated the difference between the immunopathology for a viral load trajectory representative of a deceased patient (figure 4D, arrowhead on the scale), $P_{deceased}$, and the immunopathology calculated for population parameter estimates, $P_0$, for different values of $\alpha, \beta$ or $\gamma$. While the absolute value of the immunopathology may change, if the relative values for the two cases remained robust, it would imply that our results are not sensitive to the choice of values of $\alpha, \beta$ or $\gamma$. We varied these parameters 10-fold up and down and calculated the relative difference in immunopathology for each parameter set as $\frac{P_{deceased}-P_0}{P_{deceased}}$. The default value of the relative difference was 0.893 (when $\alpha, \beta, \gamma$ had values 0.001, 0.05 and 0.5, respectively). Parameters were varied individually (figure S5), and we did not observe any remarkable alteration in the relative difference in immunopathology. Our results were thus robust to variations in $\alpha, \beta$ and $\gamma$.

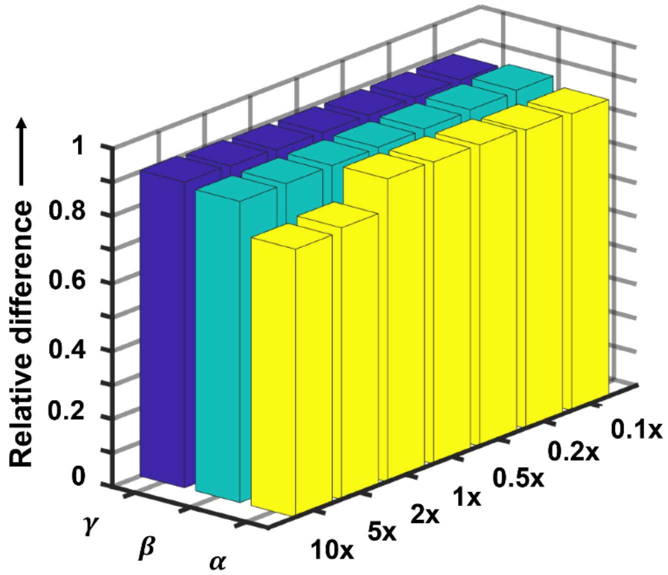


**Figure S5: Sensitivity estimates of perturbations in the parameters associated with immunopathology.** Relative difference in immunopathology between that corresponding to the population parameters and the immunopathology estimated for the deceased for different values of the parameters. Relative difference for default parameters: 0.893. $\alpha, \beta, \gamma$ were varied (from 0.1x to 10x of their default values, as indicated) individually and the corresponding relative difference calculated.

**D. Fixed points and their linear stability analysis**

We reproduce the model equations for convenience below. We ignore the equation for $D$ as it is decoupled from the rest.

$$\frac{dI}{dt} = k_1 I(1-\epsilon_I X)\left(1-\frac{I}{I_{max}}\right) - k_2 IE$$
$$\frac{dE}{dt} = \frac{k_3}{k_p+I}IE - \frac{k_4}{k_e+I}IE$$
$$\frac{dX}{dt} = k_5 I - k_6 X$$

The terms and variables are defined in the main text. We solved the equations for steady state and obtained the following fixed points:

1. $I = 0,\ E \geq 0, X = 0$
2. $I = \frac{k_6}{k_5\epsilon_I}, E = 0,\ X = \frac{1}{\epsilon_I}$
3. $I = I_{max},\ E = 0,\ X = \frac{k_5}{k_6}I_{max}$
4. $I = \frac{k_e k_3 - k_p k_4}{k_4 - k_3}, E = \frac{k_1}{k2}\left(1-\epsilon_I\frac{k_5}{k_6}I\right)\left(1-\frac{I}{I_{max}}\right), X = \frac{k_5}{k_6}I$

The stability of the fixed points depends on the signs of eigenvalues of the Jacobian matrix, $J$, of the model equations evaluated at the fixed points:

$$J = \begin{pmatrix} \frac{\partial\left(\frac{dI}{dt}\right)}{\partial I} & \frac{\partial\left(\frac{dI}{dt}\right)}{\partial E} & \frac{\partial\left(\frac{dI}{dt}\right)}{\partial X} \\ \frac{\partial\left(\frac{dE}{dt}\right)}{\partial I} & \frac{\partial\left(\frac{dE}{dt}\right)}{\partial E} & \frac{\partial\left(\frac{dE}{dt}\right)}{\partial X} \\ \frac{\partial\left(\frac{dX}{dt}\right)}{\partial I} & \frac{\partial\left(\frac{dX}{dt}\right)}{\partial E} & \frac{\partial\left(\frac{dX}{dt}\right)}{\partial X} \end{pmatrix}_{I,E,X}$$

We populated the Jacobian matrix and computed the eigenvalues. For fixed point 1, the eigenvalues were $-k_6$, 0, and $k_1 - k_2 E$. Note that the fixed point is really a line of fixed points on the E-axis (see figure S7). It follows from the eigenvalues that the fixed points with $E < \frac{k_1}{k_2}$ act as saddle points while those with $E > \frac{k_1}{k_2}$ act as marginally stable (attracting) fixed points. The latter represent clearance. When the initial value of $E = 0$, there is a stable steady state in the $E = 0$ plane (figure S1).

For fixed point 2, the eigenvalues were:

1. $\left(\frac{k_3}{k_p+\frac{k_6}{k_5\epsilon_I}} - \frac{k_4}{k_e+\frac{k_6}{k_5\epsilon_I}}\right)\frac{k_6}{k_5\epsilon_I}$
2. $k_6\left(-1+\sqrt{1+\frac{4k_1}{k_5\epsilon_I I_{max}}\left(1-\epsilon_I\frac{k_5}{k_6}I_{max}\right)}\right)$
3. $k_6\left(-1-\sqrt{1+\frac{4k_1}{k_5\epsilon_I I_{max}}\left(1-\epsilon_I\frac{k_5}{k_6}I_{max}\right)}\right)$

Depending on the parameters, which we discuss below, this fixed point could be stable or unstable.

For fixed point 3, the eigenvalues were

1. $-k_6$
2. $\frac{k_3 I_{max}}{k_p + I_{max}} - \frac{k_4 I_{max}}{k_e + I_{max}}$
3. $-k_1 \left(1 - \epsilon_I \frac{k_5}{k_6} I_{max}\right)$

Again, the fixed point is stable or unstable depending on the parameters.

Finally, for fixed point 4, the calculation of the eigenvalues explicitly was not required as we established that it was always unstable for positive values of $E$.

Fixed points 2 and 3 represented persistent infection where the CD8 T-cell response was suppressed by the virus. Importantly, their stability was independent of the stability of the clearance state. Thus, parameter combinations could be identified where clearance and persistence were both stable. The system could thus exhibit bistability. Such bistability has been proposed earlier for other viral infections, including HIV (4, 6–8). The outcomes realized would then depend on initial conditions. By analyzing parameter regimes, we identified combinations when fixed points 2 and 3 could be stable. The regimes are depicted in figure S6.

For the best-fit population parameter estimates (table S6), the system admitted a single stable steady state, fixed point 1, indicating that clearance was the only outcome realized (figure S7A). The path to clearance, however, could vary widely and depend on the initial CD8 T-cell population. A large initial effector pool could facilitate rapid clearance, in agreement with observations of such clearance facilitated by cross-reactive effector T-cells (9, 10). If the parameters are varied in a way that the strength of the immune system decreases, bistability is introduced into the system. One of the fixed points, 2 or 3, becomes stable depending on the parameter regimes. Depending on the initial conditions, trajectories can either go towards clearance or persistence (figure S7). The latter trajectories were associated with large infected cell numbers and high inflammatory cytokine levels. Such trajectories may end prematurely due to mortality because of high immunopathology, above the threshold range (see figure 4). We note that trajectories heading towards fixed point 1 may also be similarly terminated if excessive immunopathology results.

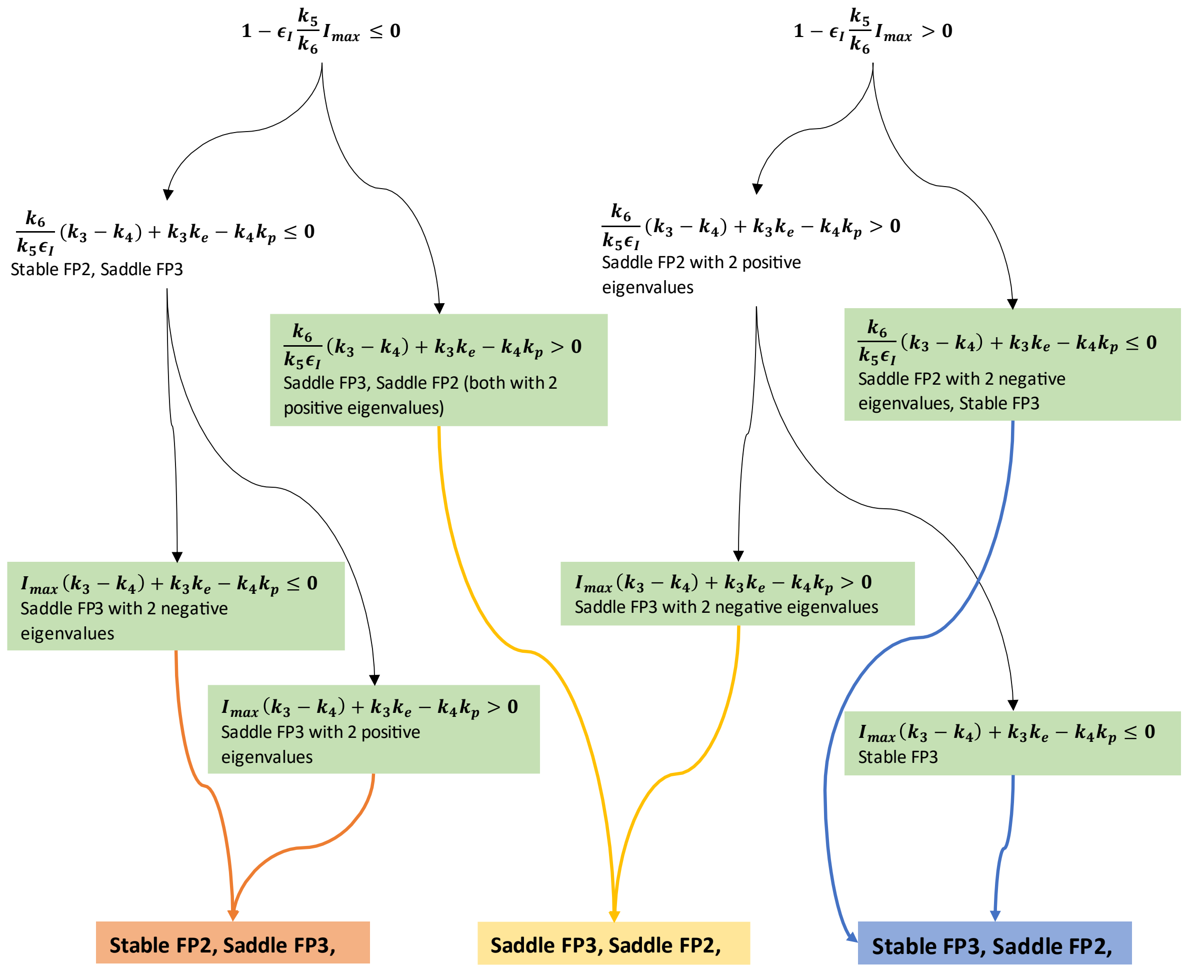


**Figure S6: Flowchart depicting parameter regimes defining the stability of fixed points 2 and 3.**

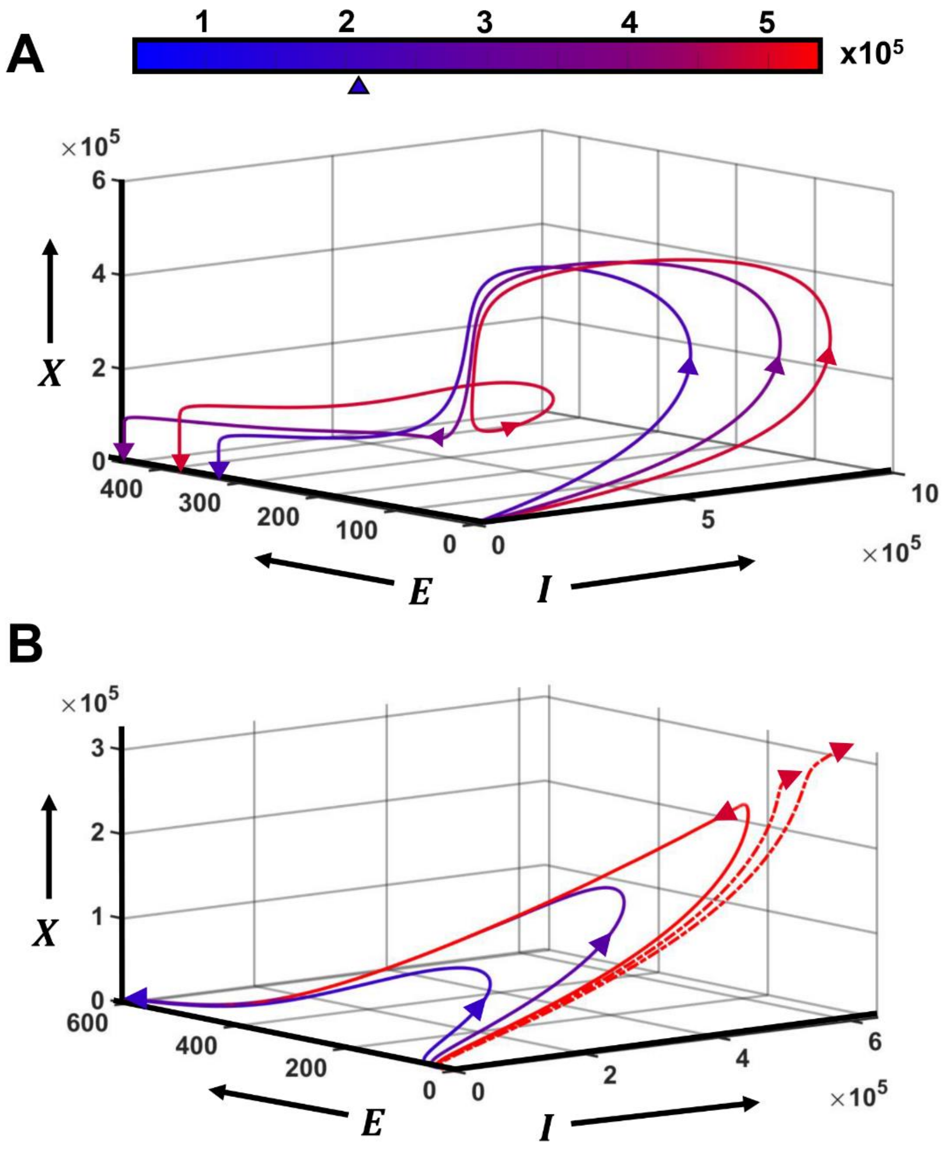


**Figure S7: Monostability and bistability, sample trajectories and associated immunopathology.** (A) Trajectories in 3D space defined by infected cells, CD8 T-cells and cytokine-mediated innate response for parameter combinations where clearance alone is a stable fixed point. Each trajectory uses different model parameter sets. The colors of the trajectories represent the immunopathology associated, defined in the scale bar above. Immunopathology corresponding to population parameter estimates is marked with an arrowhead. (B) Parameter combinations which lead to bistability with stable fixed points 1 and 3 are used. Dashed red lines are the trajectories lying on the other side of the separatrix and are headed towards fixed point 3. For these trajectories, immunopathology diverged.

**E. Effect of varying viral inoculum size**

To check the effects of the size of viral inoculum on the trajectories of the infection, we simulated our model with the estimated population parameters and varied the initial number of infected cells from 1 to 100. Apart from the timing of the peak of infection, the dynamics played out similarly in all the cases (figure S8A). The peak viral load and the immunopathology suffered were comparable in all cases (figure S8B). This is consistent with studies on macaques where infection with different inoculum sizes led to comparable disease outcomes (11). Moreover, smaller the inoculum, longer was the onset time of the rise in viral load (11).

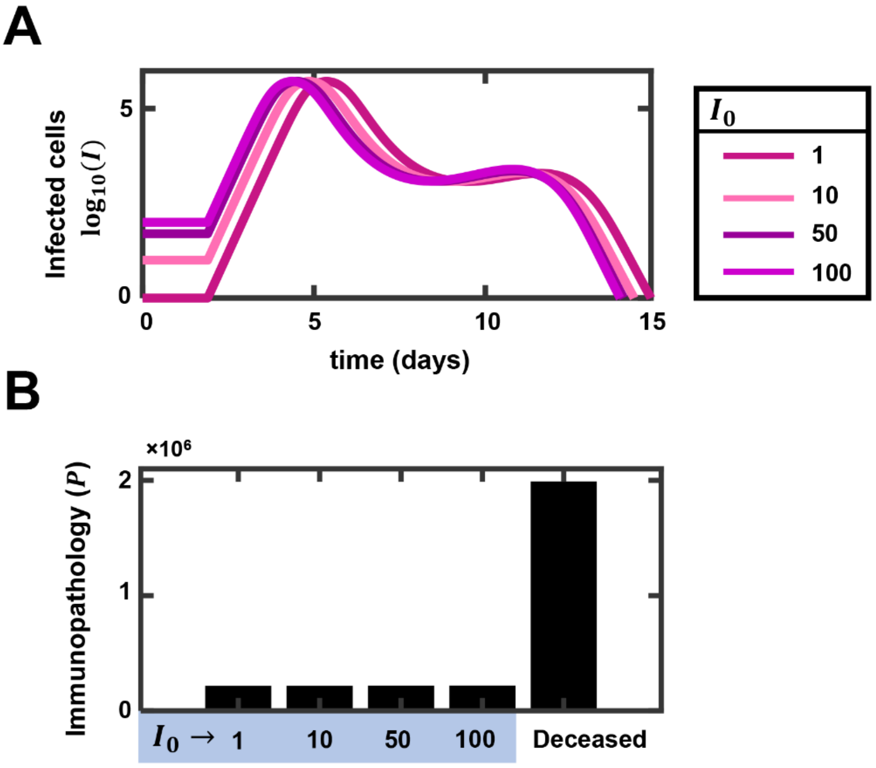


**Figure S8**: **Dependence of infection dynamics on viral inoculum size. (A)** Infected cells dynamics with varying initial infected cell pool sizes (1, 10, 50, 100 cells). **(B)** Immunopathology of the 4 trajectories compared with the calculated immunopathology for model representing deceased patient (figure 4D, arrowhead on the scale on the right).

**Table S9: Parameters of the simulations shown for non-hospitalized patient data (figure 4A).** GSA: the parameter set used as reference for global sensitivity analysis

| | Set 1 (GSA) | Set 2 | Set 3 | Set 4 |
|---|---|---|---|---|
| $k_2$ | 0.0224 | 0.0336 | 0.0224 | 0.0224 |
| $k_3$ | 0.64 | 0.64 | 0.64 | 0.64 |
| $k_p$ | 548.25 | 328.95 | 36.55 | 493.43 |
| $E_0$ | 1.26 | 1.26 | 1.26 | 2.52 |
| $k_5$ | 10 | 4 | 12 | 3 |
| $k_6$ | 0.24 | 0.24 | 0.24 | 0.24 |
| $\varepsilon_I$ | 3.16e-05 | 3.16e-05 | 2.53e-05 | 3.48e-05 |
| $\tau$ | 1.4 | 1.9 | 3.9 | 5.9 |

**Table S10: Parameters of the simulations shown for moderate patient data (figure 4B).** GSA: the parameter set used as reference for global sensitivity analysis

| | Set 1 | Set 2 | Set 3 (GSA) | Set 4 |
|---|---|---|---|---|
| $k_2$ | 0.0224 | 0.0224 | 0.0448 | 0.0224 |
| $k_3$ | 0.64 | 0.64 | 0.64 | 0.4267 |
| $k_p$ | 3.29e+03 | 2193 | 2193 | 2.74e+03 |
| $E_0$ | 1.26 | 1.26 | 1.26 | 1.26 |
| $k_5$ | 3 | 3 | 5 | 6 |
| $k_6$ | 0.24 | 0.24 | 0.24 | 0.24 |
| $\varepsilon_I$ | 1.58e-06 | 1.58e-06 | 1.58e-06 | 1.58e-06 |
| $\tau$ | 1.4 | 2.9 | 3.9 | 4.9 |

**Table S11: Parameters of the simulations shown for severe patient data (figure 4C).** GSA: the parameter set used as reference for global sensitivity analysis

| | Set 1 | Set 2 | Set 3 (GSA) | Set 4 |
|---|---|---|---|---|
| $k_2$ | 0.1568 | 0.0269 | 0.336 | 0.3629 |
| $k_3$ | 0.5953 | 0.5333 | 0.768 | 0.6214 |
| $k_p$ | 548.25 | 1.645e+03 | 3.29e+04 | 7.676e+03 |
| $E_0$ | 1.26 | 1.26 | 0.63 | 1.26 |
| $k_5$ | 1 | 1 | 1 | 3 |
| $k_6$ | 0.24 | 0.24 | 0.2526 | 0.24/.95 |
| $\varepsilon_I$ | 6.32e-07 | 1.053e-06 | 5.267e-07 | 3.95e-07 |
| $\tau$ | 1.4 | 2.4 | 1.9 | 3.9 |

**Table S12: Parameters of the simulations shown for deceased patient data (figure 4D).** GSA: the parameter set used as reference for global sensitivity analysis

| | Set 1 | Set 2 | Set 3 | Set 4 (GSA) |
|---|---|---|---|---|
| $k_2$ | 0.0336 | 0.0403 | 0.0403 | 0.0403 |
| $k_3$ | 0.64 | 0.64 | 0.64 | 0.64 |
| $k_p$ | 10965 | 5.483e+03 | 10965 | 21930 |
| $E_0$ | 1.26 | 1.26 | 1.26 | 1.26 |
| $k_5$ | 1 | 1 | 1 | 1 |
| $k_6$ | 0.24 | 0.24 | 0.24 | 0.24 |
| $\varepsilon_I$ | 7.9e-07 | 5.27e-07 | 7.9e-07 | 7.9e-07 |
| $\tau$ | 1.9 | 4.9 | 3.4 | 1.4 |

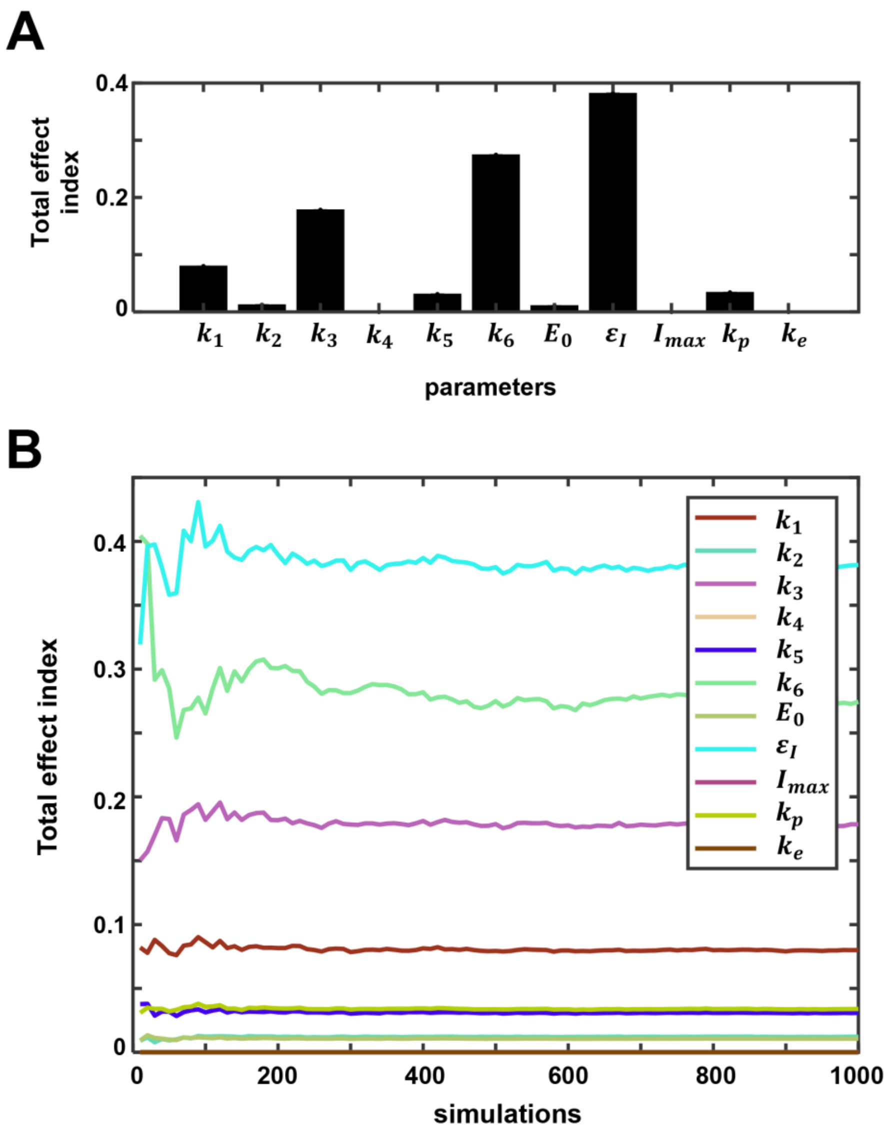


**Figure S9: Global sensitivity analysis of model parameters corresponding to non-hospitalized patients.** (A) A variance-based global sensitivity analysis on the immunopathology output of the model was performed by perturbing the parameters of the model within ±5% of the nominal value, using a Monte Carlo simulation approach. The results were summarized as the total effect indices. (B) Convergence analysis of the total effect indices for all parameters plotted in (A).

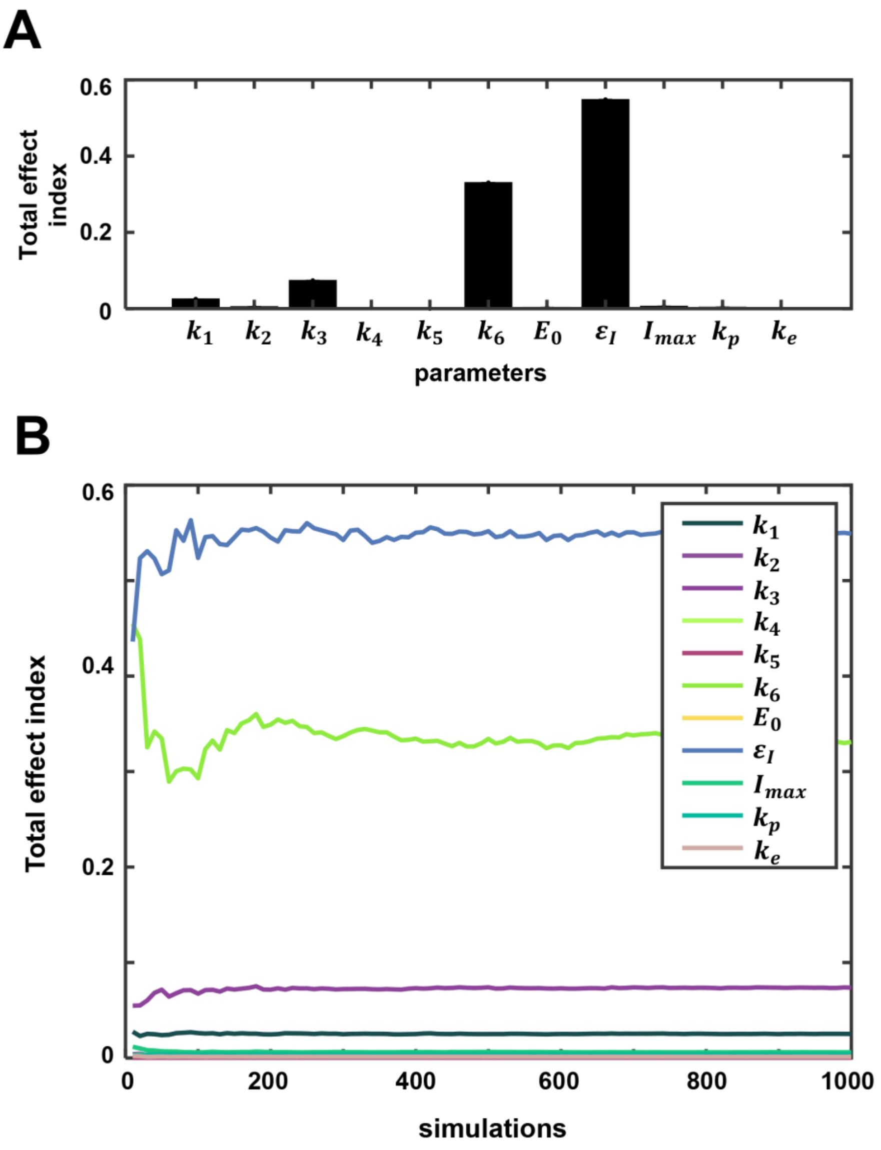


**Figure S10: Global sensitivity analysis of model parameters corresponding to patients with moderate symptoms.** (A) A variance-based global sensitivity analysis on the immunopathology output of the model was performed by perturbing the parameters of the model within ±5% of the nominal value, using a Monte Carlo simulation approach. The results were summarized as the total effect indices. (B) Convergence analysis of the total effect indices for all parameters plotted in (A).

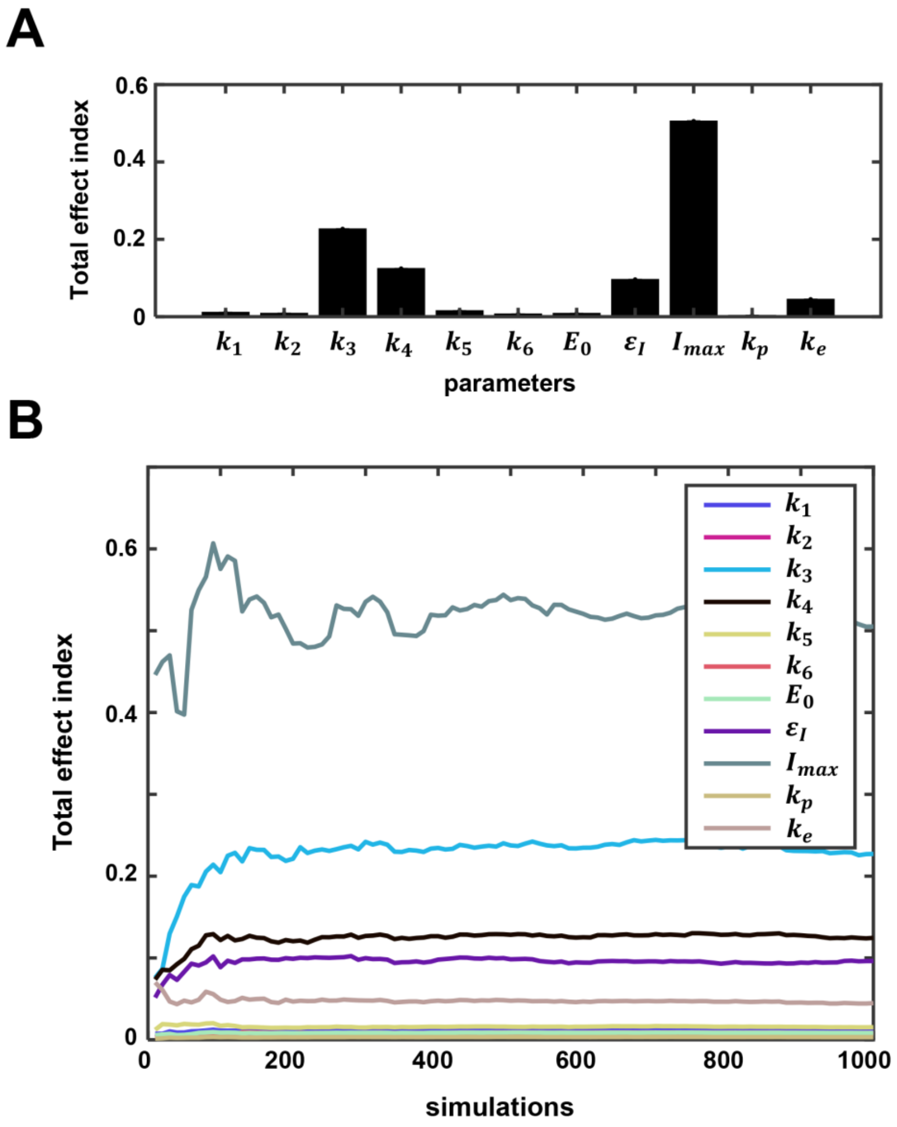


**Figure S11: Global sensitivity analysis of model parameters corresponding to patients with severe symptoms.** (A) A variance-based global sensitivity analysis on the immunopathology output of the model was performed by perturbing the parameters of the model within ±5% of the nominal value, using a Monte Carlo simulation approach. The results were summarized as the total effect indices. (B) Convergence analysis of the total effect indices for all parameters plotted in (A).

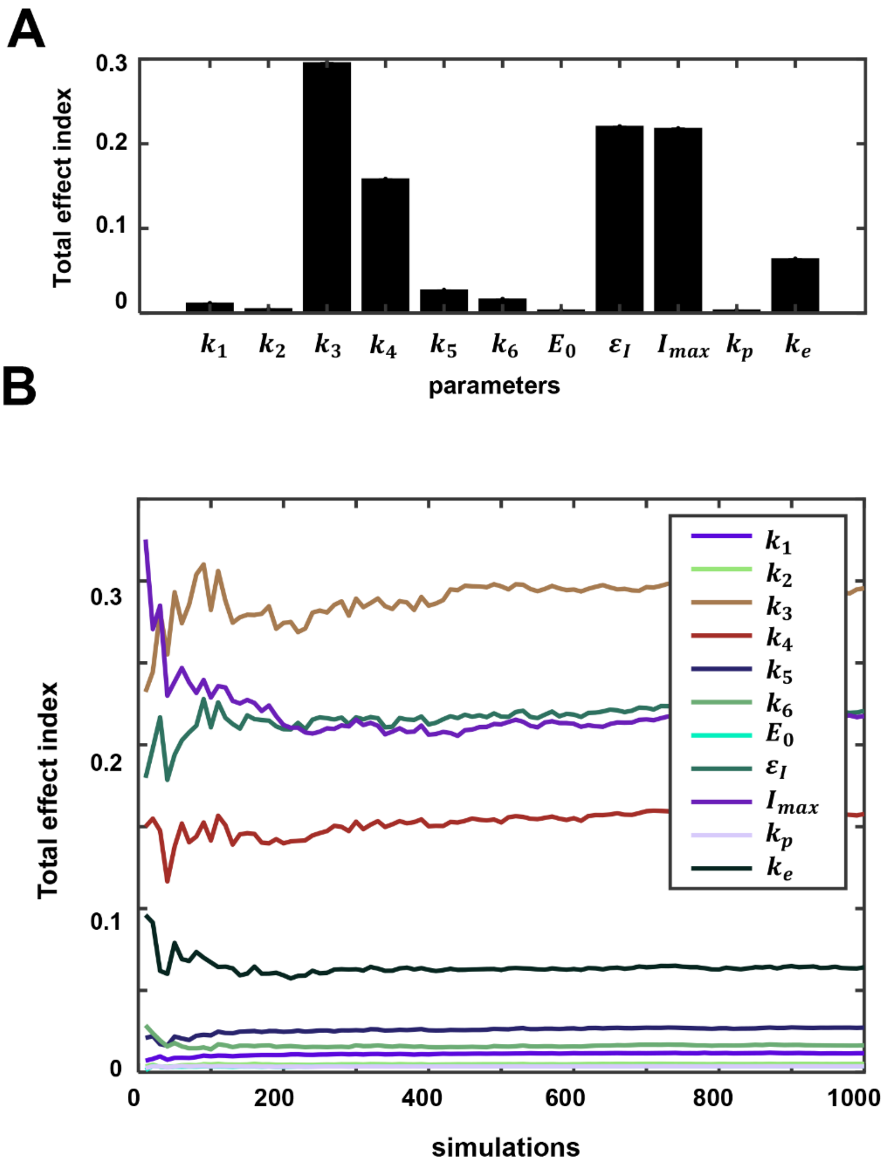


**Figure S12: Global sensitivity analysis of model parameters corresponding to deceased patients.** (A) A variance-based global sensitivity analysis on the immunopathology output of the model was performed by perturbing the parameters of the model within ±5% of the nominal value, using a Monte Carlo simulation approach. The results were summarized as the total effect indices. (B) Convergence analysis of the total effect indices for all parameters plotted in (A).
